# Distinct metabolomic and lipidomic profiles associated with cirrhosis after HCV cure in people with HIV: findings at one and five years

**DOI:** 10.64898/2026.03.24.26349149

**Authors:** Ana Virseda-Berdices, Belen Requena, Juan Berenguer, Juan Gónzalez-García, Carolina Gonzalez-Riano, Raquel Behar-Lagares, Cristina Díez, Victor Hontañón, Amanda Fernández-Rodríguez, Coral Barbas, Rubén Martín-Escolano, Salvador Resino, María Ángeles Jiménez-Sousa, the Marathon Study Group

## Abstract

**Background & Aims:** People with HIV (PWH) who achieve hepatitis C virus (HCV) cure may retain persistent metabolic alterations, particularly those with advanced fibrosis or cirrhosis. This study aimed to characterize plasma metabolomic and lipidomic profiles associated with cirrhosis in PWH at one and five years post-HCV therapy.

**Methods:** Two cross-sectional studies evaluated PWH one (n=48) and five (n=30) years post-HCV therapy. Cirrhosis was defined as a liver stiffness measurement (LSM)≥12.5 kPa. Metabolomics and lipidomics were performed using capillary electrophoresis-mass spectrometry (CE-MS) and liquid chromatography-mass spectrometry (LC-MS), respectively. Data were analyzed using orthogonal partial least squares discriminant analysis (OPLS-DA) and generalized linear models (GLM), adjusting for relevant covariates.

**Results:** At one and five years, 32 (66.7%) and 10 (33.3%) participants, respectively, had cirrhosis. OPLS-DA identified 235 and 229 metabolites with variable importance in projection (VIP)scores >1. At one year, cirrhosis was associated with elevated levels of glycerophospholipids, sphingomyelins, and amino acids, and lower levels of triglycerides. At five years, cirrhotic PWH exhibited higher levels of glycerophospholipids and acyl-carnitines, together with lower levels of triglycerides and amino acids.

**Conclusions:** PWH with cirrhosis post-HCV cure exhibits a persistently altered metabolic profile stable for five years, suggesting ongoing liver disease progression. These findings underscore the need for continued long-term monitoring of this population.

## Introduction

Sustained virologic response (SVR), whether achieved through interferon (IFN)-based regimens or all-oral direct-acting antivirals (DAAs), profoundly alters the natural history of hepatitis C virus (HCV) infection ^1^. SVR reduces the risk of progression to cirrhosis, hepatic decompensation, hepatocellular carcinoma (HCC), and both liver-related and all-cause mortality ^2, 3^. Beyond hepatic outcomes, SVR is also associated with a lower incidence of extrahepatic manifestations, including cardiovascular events as well as renal and metabolic complications ^2, 3^.

Nevertheless, a subset of patients with advanced fibrosis or cirrhosis continue to experience disease progression and remain at residual risk of HCC ^4^. This risk is particularly pronounced in the presence of additional liver-damaging factors, such as excessive alcohol consumption ^5^, metabolic dysfunction–associated steatotic liver disease (MASLD) ^6^, or coinfection with hepatotropic viruses, including hepatitis B virus (HBV) with or without hepatitis D virus (HDV) ^7^. These findings highlight the importance of continued surveillance in patients with advanced liver disease, even after HCV clearance ^8^.

Recent advances in metabolomics and lipidomics offer powerful tools to unravel the molecular mechanisms underlying liver disease pathophysiology. Comprehensive profiling of metabolites and lipids can reveal perturbations in pathways related to inflammation, fibrogenesis, and hepatic dysfunction, and may enable the identification of biomarkers predictive of disease progression and clinical outcomes ^9, 10^. These metabolic disorders have been found in several viral infections ^11^, including HIV. People with HIV (PWH) represent a particularly vulnerable population. HIV infection disrupts multiple metabolic pathways, and although antiretroviral therapy (ART) partially restores them, persistent alterations and chronic immune activation may contribute to long-term hepatic and metabolic complications ^12, 13^.

This study aims to investigate the metabolomic and lipidomic profiles of PWH who continue to present with cirrhosis despite achieving HCV cure. By characterizing molecular alterations in this high-risk population, we aim to shed light on the residual mechanisms driving liver disease progression after viral eradication.

## Methods

### Study subjects

We conducted two cross-sectional studies including PWH on long-term suppressive ART: 48 participants assessed one year, and 30 participants assessed five years after successful completion of HCV therapy, with 24 individuals assessed at both time points. HCV treatment regimens included either IFN-free DAAs or IFN-based therapy (peg-IFN-α/ribavirin or peg-IFN-α/ribavirin/DAAs). Participants were recruited from 10 centers across Spain between 2012 and 2021 (Marathon Study Group; **Appendix A**).

SVR was defined as an undetectable HCV-RNA load 12-24 weeks after treatment completion. All patients were receiving stable ART for at least six months and had an undetectable plasma HIV viral load (<50 copies/mL). Individuals with acute hepatitis C, hepatitis B virus (HBV) infection, or a prior history of liver decompensation were excluded.

This study was approved by the Research Ethics Committee of the Institute of Health Carlos III (CEI PI 72_2021) and conducted in accordance with the Declaration of Helsinki. All participants gave their written informed consent before enrollment.

### Clinical data and samples

Epidemiological and clinical data were retrospectively collected using a secure online platform, and subsequently verified against patient medical records to ensure accuracy and consistency. LSM were performed by trained operators using transient elastography, as previously described ^14^.

Peripheral blood samples were collected in ethylenediaminetetraacetic acid (EDTA) tubes, and plasma was isolated by Ficoll-Paque density gradient centrifugation. Samples were stored at -80°C in the Spanish HIV HGM Biobank until shipment for analysis.

### Primary study variable

The primary study variable was the presence of cirrhosis, defined as LSM≥12.5 kPa. This threshold was selected because it is a widely validated and commonly used cutoff for the non-invasive diagnosis of cirrhosis ^15^. Data were analyzed separately for the cross-sectional assessment performed at one year and five years after completion of HCV therapy.

### Non-targeted metabolomics analysis

A detailed description of metabolite extraction, reagents and standards, quality control (QC), procedures, analytical conditions, and metabolite annotation is provided in **Supplementary Data 1**.

### Metabolites extraction and quality assurance

Inactivated plasma samples were prepared at the Centro de Metabolómica y Bioanálisis, CEMBIO (Madrid, Spain). For CE-MS analysis, the procedure involved drying on the SpeedVac Concentrator (Thermo Fisher Scientific, Waltham, MA, USA), a deproteinization process, and transference to a chromacol vial. For LC-MS, processed plasma samples were extracted using a procedure that included methanol and methyl-tert-butyl ether (MeOH:MTBE (1:1, v/v))^16^.

For quality management assurance and blank samples, each plasma sample was polled in equal quantities to create QC samples. These QC samples were processed in the same way as the other samples. To detect common contaminations, two blank samples were prepared and processed using the same lipid extraction method as the other samples. The analytical sequence started and ended with injections of these samples.

### Capillary Electrophoresis Analysis (CE-MS)

We carried out an untargeted metabolomics-based approach to capture the diverse spectrum of plasma metabolites. Agilent CE 7100 coupled with an Agilent 6224 time-of-flight Mass Spectrometer (TOF-MS) analyzer was used to analyze the samples. We used a fused-silica capillary (Agilent Technologies; total length 100 cm x 50 μm i.d. x 360 µm) for CE-MS separation analysis in positive ionization mode. ESI (+) positive polarity mode was used in mass spectrometer (MS). Finally, CE-MS system was operated using MassHunter Workstation 6200 series TOF version B.09.00 c(B9044.0) acquisition software.

### Lipidomics Analysis (LC-MS)

Agilent 1290 Infinity II Ultra-High-Performance Liquid-Chromatography (UHPLC) system coupled to an Agilent 6546 Quadrupole Time-of-Flight (QTOF) Mass Spectrometer (MS) equipped with dual Agilent Jet Stream (AJS) Electrospray (ESI) ion source (separate runs in positive and negative ESI modes) was used to analyze the samples. We used MassHunter Workstation Software LC-MS Data Acquisition v B.09.00 (Agilent Technologies, Waldrobonn, Germany) to obtain MS/MS information for plasma lipidome.

### Data reprocessing

Hybrid strategies were followed for the reprocessing of the raw data files, consisted of using an extensive compound database, both lipidomics and metabolomics data, respectively, which was imported into the Agilent MassHunter Profinder software (B.10.0.2, Agilent Technologies, Santa Clara, CA, USA) to perform the time alignment and feature extraction using the "Batch Targeted Feature Extraction" mode. For CE-MS data reprocessing we employed an in-house library ^17^, included in the CEU Mass Mediator (CMM) online tool ^18^.

To construct the lipidomic database, we performed a comprehensive lipid annotation process using four bioinformatic tools. This process incorporates a bottom-up strategy (LipidHunter), spectra matching (Lipid Annotator, MS-Dial), and the fragment intensity rules (LipidMS) ^19^. Then, false software annotations and redundant data were removed following the review of tentative annotations supplied by the software tools. To obtain a final list of lipids, we used the assistance of the CEU Mass Mediator (CMM) ^18^ and the manual inspection of the MS and MS/MS spectra data. **Supplementary Data 1** shows full description of the software parameters. The integration of multiple complementary tools enhanced the accuracy and reliability of the lipid annotation.

Databases included the migration time (MT), monoisotopic exact mass, and molecular formula. Only features that were present in at least 50% of the samples within a single sample group, were taken into consideration as a last post-processing filter.

For CE-MS metabolomics data, the methionine sulfone IS, which was added in the initial preparation phase, was used for normalization. Lipidomic data normalization was carried out using the Kuligowski transformation ^20^. Then, we applied a threshold of 30%, based on the coefficient of variation (CV) within the QC samples, to select metabolites and lipid species.

### Statistical analysis

Statistical analysis was performed using MetaboAnalyst 6.0 software (http://www.metaboanalyst.ca/) and R software version 4.3.1 (R Foundation for Statistical Computing, Vienna, Austria).

For descriptive analysis, categorical variables were expressed as absolute counts (percentages), and continuous variables were shown as median and interquartile range (IQR). Chi-square and Mann-Whitney U tests were used to compare categorical and quantitative variables, respectively.

For the multivariate analysis, firstly, metabolomic variables were log-transformed (log_2_) and auto-scaled. Next, we performed a supervised multivariate analysis by orthogonal partial least squares discriminant analysis (OPLS-DA), which models simultaneously all features. Variable importance in projection (VIP) scores were calculated for each feature and model validity was assessed using a 1,000-permutation test.

For the association analysis, Generalized Linear Models (GLM) with gamma distribution was performed to evaluate the association between individual metabolomic features (dependent variable) and cirrhosis (LSM≥12.5 KPa) (independent variable) one year and five years after successful completion of HCV therapy. The models were adjusted for relevant covariates (age, gender, BMI, and HCV treatment) using a forward stepwise selection method based on the lowest Akaike Information Criterion (AIC). Results were reported as the adjusted arithmetic means ratio (aAMR) and corresponding p-values. Multiple testing correction was performed using the false discovery rate (FDR) according to Benjamini and Hochberg procedure. Statistical significance was defined as p-value <0.05 and an FDR-adjusted p-value (q-value) <0.2.

### Lipid pathways

Functional associations of metabolites significatively associated to cirrhosis were analyzed and visualized by Lipid Network Explorer platform (LINEX) ^21^. The logarithm of fold change (LFC) was used to quantify differences in lipid abundances and to visualize variation patterns across groups.

## Results

### Patient characteristics

Clinical and epidemiological characteristics of the 48 PWH assessed one year after completion HCV treatment and of the 30 individuals evaluated five years post-treatment are summarized **Table 1**.

**Table 1.**
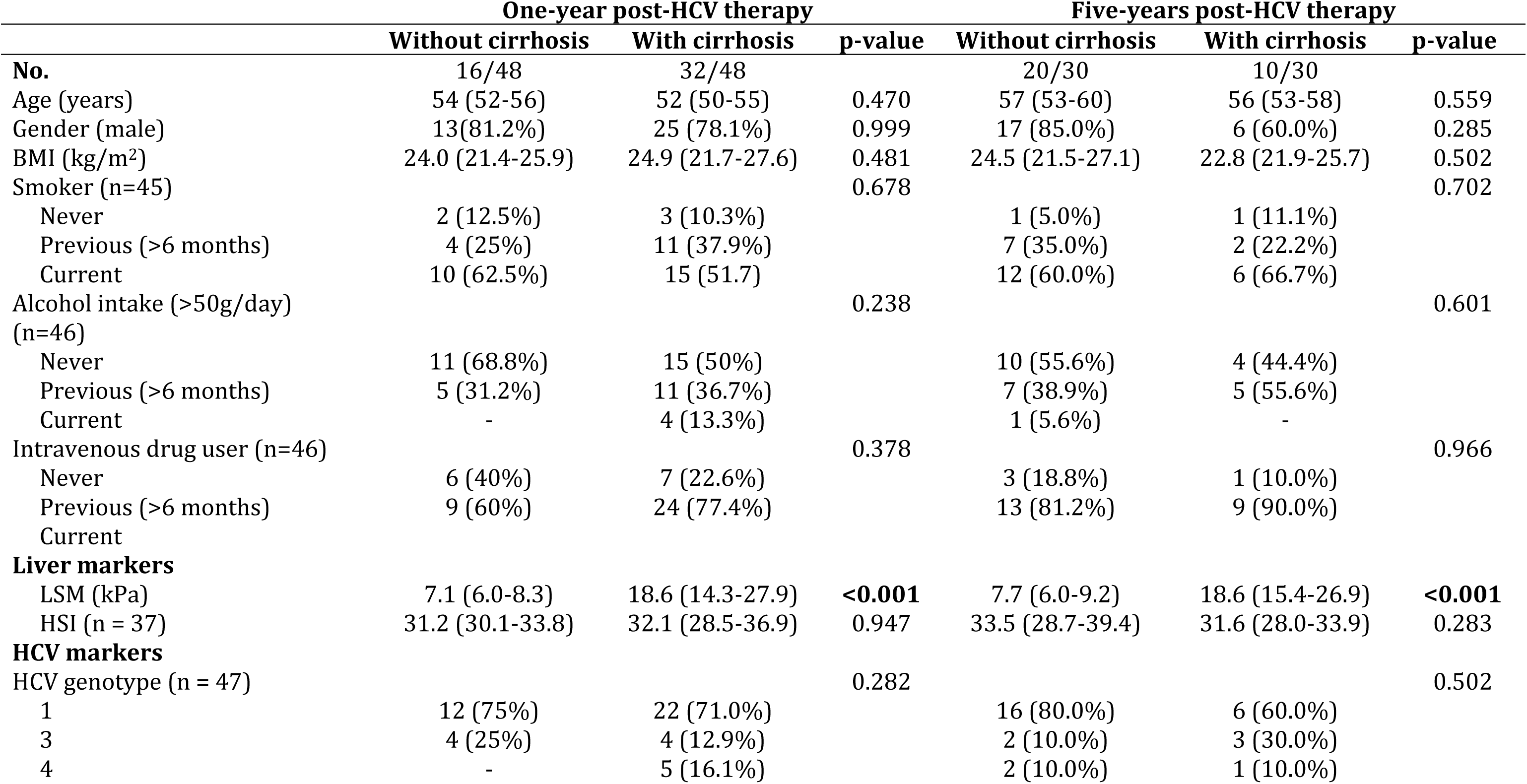
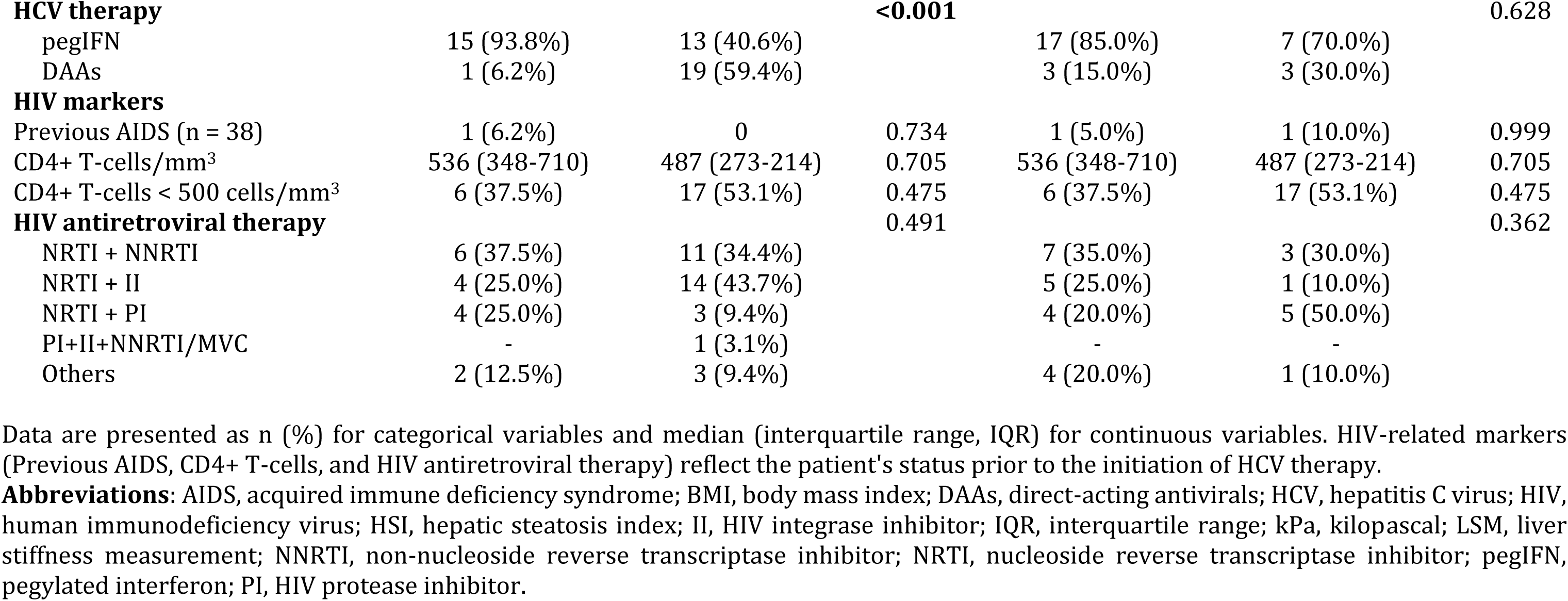
Epidemiological and clinical characteristics of people with HIV (PWH) cured of HCV, stratified by cirrhosis status one and five years post-HCV therapy.

At one-year time point, 32 individuals (67%) had cirrhosis. Significant differences between patients with and without cirrhosis were observed in LSM and type of HCV treatment (p<0.001 for both). At the five-year assessment, 10 individuals (33%) had cirrhosis, with significant differences between groups in both LSM and FIB-4 score (p<0.001 for both).

The evolution of LSM from baseline to one- and five-years is displayed in **Supplementary Data 2**. In addition, significant differences were found in platelet counts, international normalized ratio (INR), and aspartate aminotransferase (AST) levels at one year, as well as in hemoglobin levels at five years after HCV therapy (**Supplementary Data 3, Supplementary Data 4**).

### Metabolite detection in plasma samples

A total of 646 plasma metabolites were detected across the different platforms: 80 by CE-MS, 440 by LC-MS ESI+ and 126 by LC-MS ESI-. In the CE-MS analysis, most identified metabolites were amino acids (33.8%) and amino acid derivatives (45.0%). Based on the LIPID MAPS Structure Database (LMSD), lipid species were classified into four main categories, as detailed in **Supplementary Data 5**. In total, 112 glycerolipids (GL) (19.8%), 286 glycerolphospolipids (GP) (50.6%), 92 sphingolipids (SP) (16.3%), and 74 fatty acyls (13.1%) were identified.

### Metabolic profiling associated with cirrhosis one year after post-HCV therapy

OPLS-DA models effectively discriminated patients according to cirrhosis status, identifying 235 plasma metabolites with VIP scores >1 (**Figure 1A**, **Figure 2A**). Subsequent GLM analysis identified 80 metabolomic features significantly associated with cirrhosis. Among these, 13 features were found at lower proportion (AMR <1), and 67 at higher proportion (AMR ≥1) in PWH with cirrhosis. As shown in **Figure 3A**, significant metabolites detected by CE-MS included five amino acids and two derivatives. Lipids, identified by LC-MS were distributed across four categories: fatty acyls (2 compounds), SP (10), GP (54), and GL (4). Detailed association data for significant metabolites and lipids are shown in **Supplementary Data 6.**

**Figure 1.**
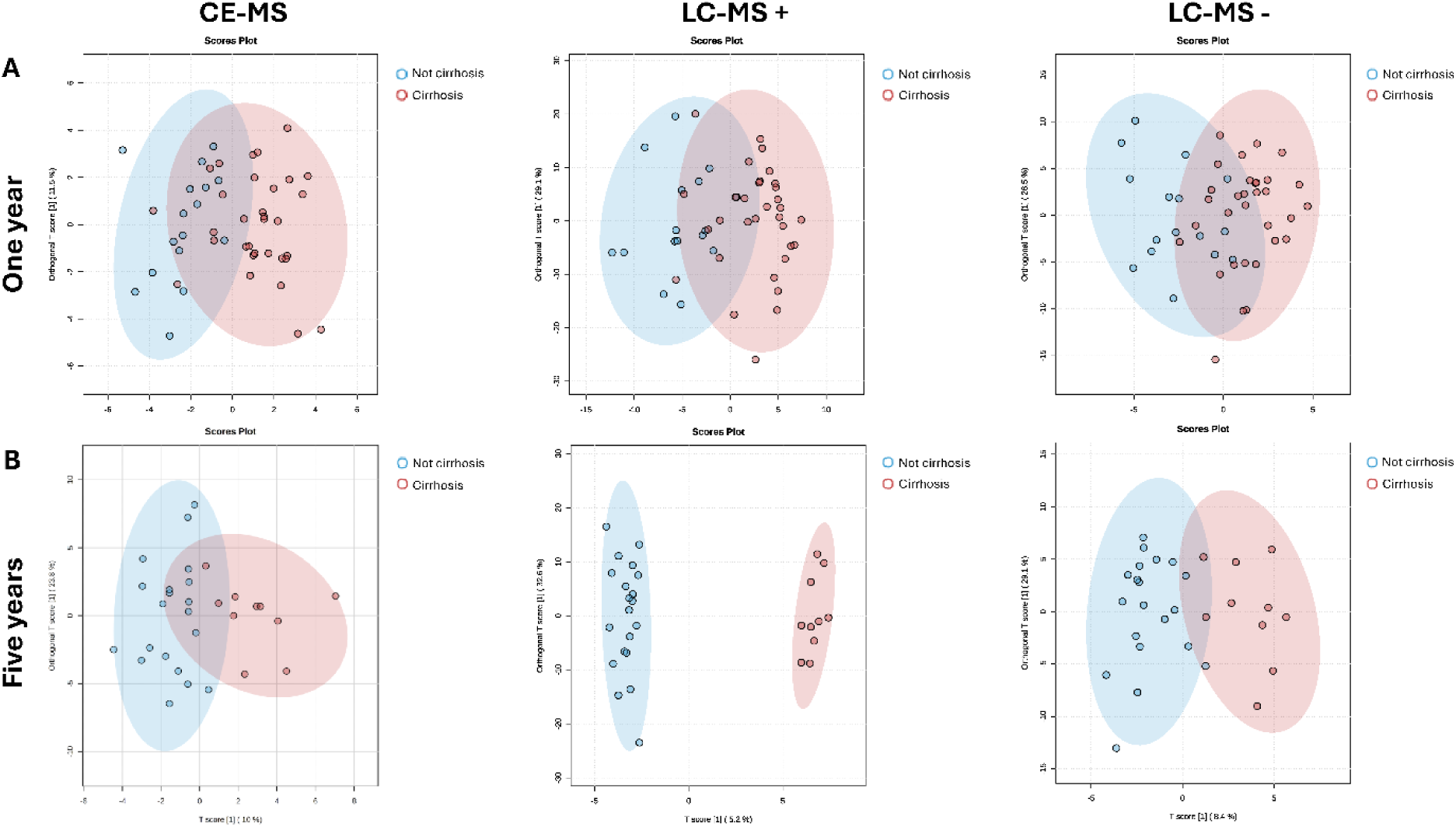
Orthogonal Partial Least Square Discriminant Analysis (OPLSDA) scores plots discriminating between PWH with and without cirrhosis. The plots show results from three different analytical platforms: (left) Capillary Electrophoresis-Mass Spectrometry (CE-MS), (center) Liquid Chromatography-Mass Spectrometry in positive ion mode (LC-MS+), and (right) LC-MS in negative ion mode (LC-MS-). Panels show analysis at (**A**) one year and (**B**) five years post-HCV therapy. **Abbreviations**: DG, diglyceride; FA, fatty acid; FAHFA, fatty acyl ester of hydroxy fatty acid; LPC, lysophosphatidylcholine; LPE, lysophosphatidylethanolamine; MG, monoglyceride; PA, phosphatidic acid; PC, phosphatidylcholine; PE, phosphatidylethanolamine; PI, phosphatidylinositol; PS, phosphatidylserine; SM, sphingomyelin; TG, triglyceride.

**Figure 2.**
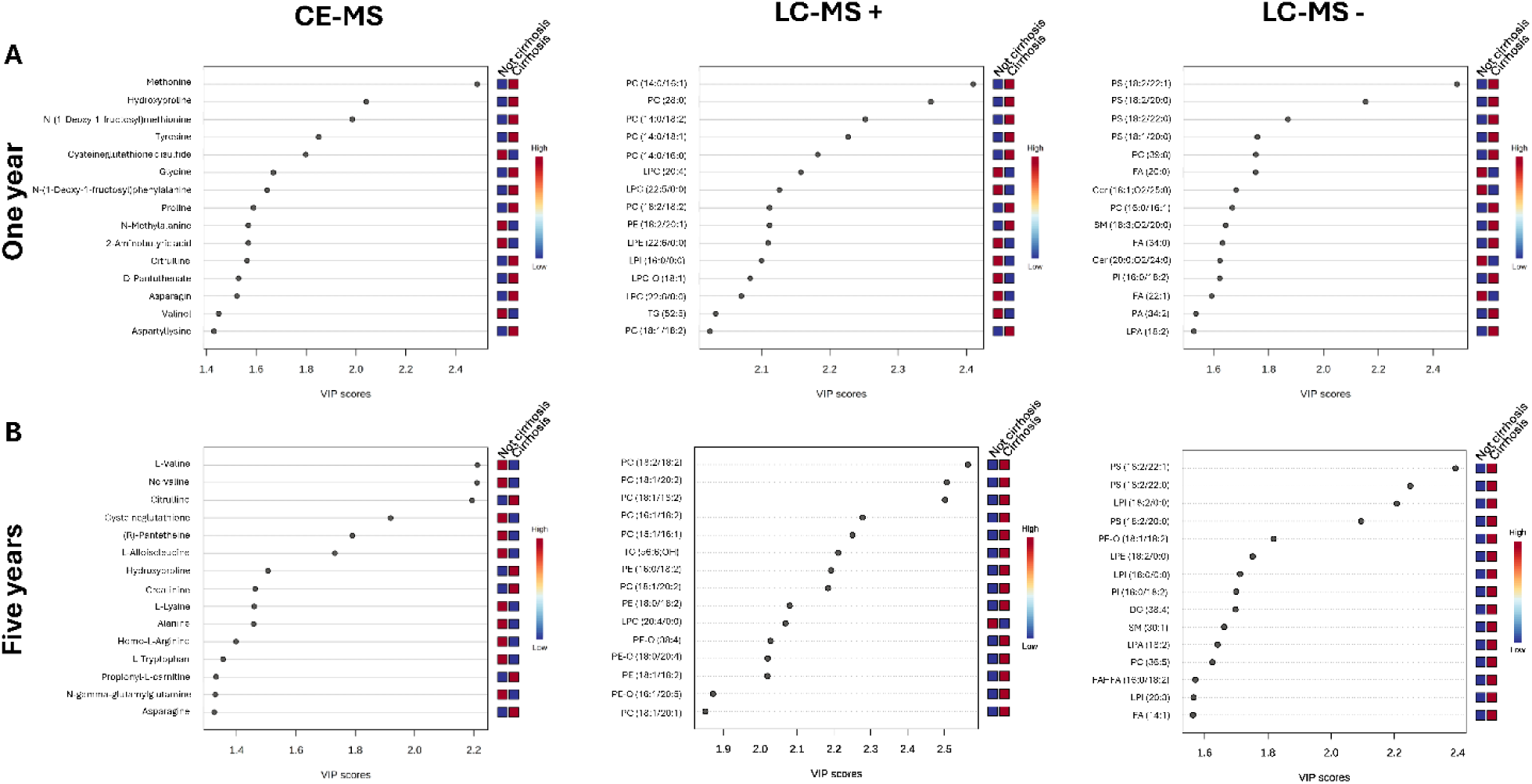
**Top 15 metabolites with the highest variable’s importance (VIP) scores (≥1) from OPLSDA models**. The plots show the most influential features discriminating between PWH with and without cirrhosis at (**A**) one year and (**B**) five years post-HCV therapy, separated by analytical platform. The color scale indicates the relative abundance of each metabolite: red bars indicate higher abundance, and blue bars indicate lower abundance. **Abbreviations**: DG, diglyceride; FA, fatty acid; FAHFA, fatty acyl ester of hydroxy fatty acid; LPC, lysophosphatidylcholine; LPE, lysophosphatidylethanolamine; MG, monoglyceride; PA, phosphatidic acid; PC, phosphatidylcholine; PE, phosphatidylethanolamine; PI, phosphatidylinositol; PS, phosphatidylserine; SM, sphingomyelin; TG, triglyceride.

**Figure 3.**
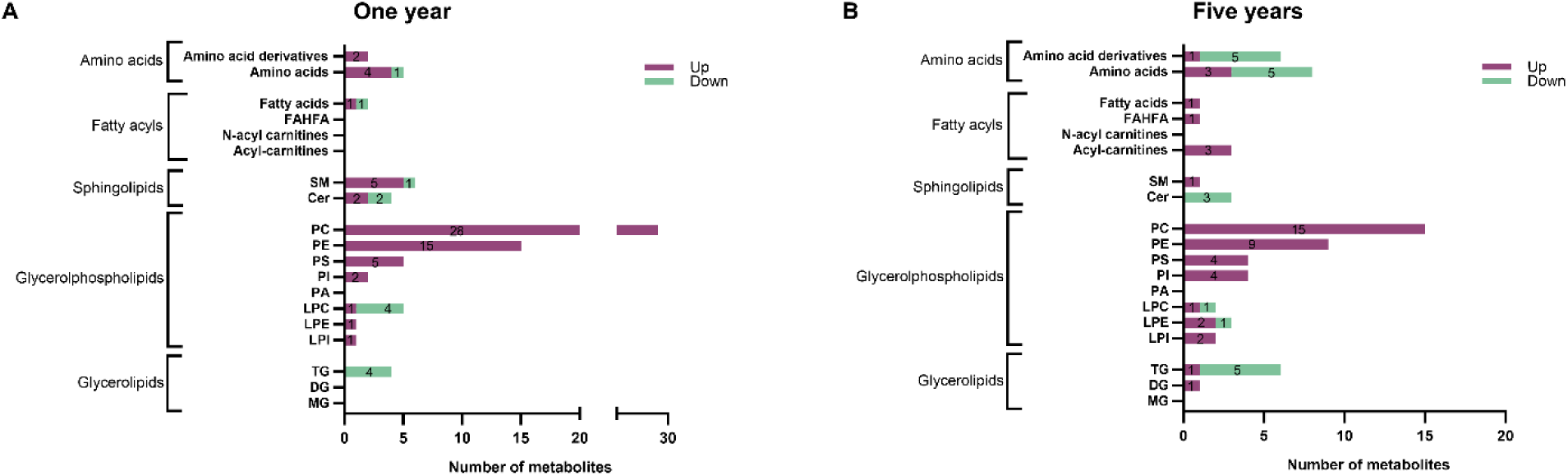
**Significantly altered metabolites in individuals with cirrhosis**. The bar charts display the count of metabolites that were significantly upregulated (purple) or downregulated (green) when comparing PWH with high vs. low LSM at (**A**) one year and (**B**) five years post-HCV therapy. Metabolites are grouped by class. **Statistical analysis**: Significance was determined using Generalized Linear Models (GLM), adjusted for age, gender, BMI, and HCV treatment. **Abbreviations**: Cer, ceramides; DG, diglyceride; FAHFA, fatty acyl ester of hydroxy fatty acid; LPC, lysophosphatidylcholine; LPE, lysophosphatidylethanolamine; LPI, lysophosphatidylinositol; MG, monoglyceride; PC, phosphatidylcholine; PE, phosphatidylethanolamine; PI, phosphatidylinositol; PS, phosphatidylserine; SM, sphingomyelin; TG, triglyceride.

Higher plasma levels of amino acids and their derivates were observed in PWH with cirrhosis (**Figure 3A**). Within the lipidomic profile, phosphatidylcholines (PC) and phosphatidylethanolamines (PE) were the most prevalent lipid species identified, with chain length modification and desaturation being the most common alterations observed (**Figure 4A**).

**Figure 4.**
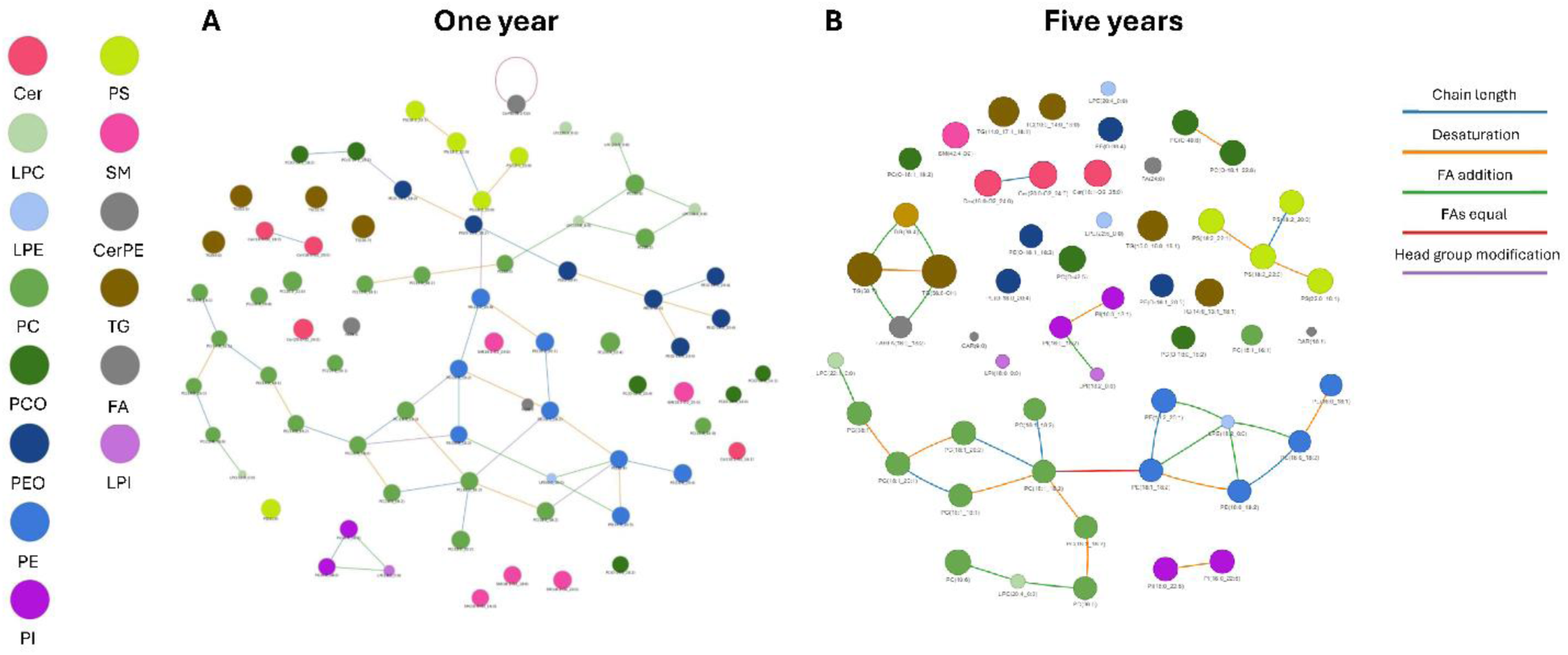
**Lipid network analysis of metabolites associated with cirrhosis**. The networks illustrate the biochemical relationships between significantly altered lipids in PWH at (**A**) one year and (**B**) five years post-HCV therapy. Nodes represent individual lipid species (colored by lipid class), and edges represent biochemical reactions connecting them (colored by reaction type). **Abbreviations**: Cer, ceramide; CerPE, ceramide-phosphate-ethanolamine; DG, diglyceride; FA, fatty acid; FAHFA, fatty acyl ester of hydroxy fatty acid; LPC, lysophosphatidylcholine; LPE, lysophosphatidylethanolamine; LPI, lysophosphatidylinositol; MG, monoglyceride; PA, phosphatidic acid; PC, phosphatidylcholine; PCO, ether-linked phosphatidylcholine; PE, phosphatidylethanolamine; PEO, ether-linked phosphatidylethanolamine; PI, phosphatidylinositol; PS, phosphatidylserine; SM, sphingomyelin; TG, triglyceride.

Regarding chemical composition, most PC species were polyunsaturated (64.3%), followed by monounsaturated (21.4%) and saturated (14.3%). In contrast, all PE and phosphatidylinositol (PI) species, as well as all but one of phosphatidylserine (PS) species, were polyunsaturated (**Supplementary Data 7A**).

### Metabolic profiling associated with cirrhosis five years post-HCV therapy

OPLS-DA models adequately separated patients according to cirrhosis status, identifying 229 plasma metabolites with VIP scores >1 (**Figure 1B**, **Figure 2B**). Of these, 69 features were significantly associated with cirrhosis, with 20 found at lower levels (AMR<1), and 49 at higher levels (AMR≥1). The distribution of significant metabolites and lipids is shown in **Figure 3B**, with full association data provided in **Supplementary Data 8.**

At five years, individuals with cirrhosis exhibited lower plasma levels of amino acids and their derivatives (**Figure 3B**). Consistent with the one-year findings, PC and PE were the predominant lipid species associated with cirrhosis. The most frequent lipid modifications included chain length modifications, desaturation, and fatty acid (FA) additions (**Figure 4B**).

In terms of glycerophospholipids composition, a marked shift was observed in PC species compared to one-year time point: saturated PCs were no longer detected, while the proportions of polyunsaturated (73.3%) and monounsaturated (26.7%) species increased. No relevant compositional changes were found for PE, PS and PI (**Supplementary Data 7B**).

## Discussion

This study is, to our knowledge, the first to longitudinally characterize the plasma metabolic profile in PWH with cirrhosis after HCV cure. We found a persistent metabolic dysregulation up to five years post-HCV eradication, marked by elevated glycerophospholipids (PC, PE, PS and PI) and reduced amino acids and triglycerides. Over time, we also observed dynamic shifts in lysophospholipids (LPC, LPE and LPI), fatty acyls, and sphingolipids (SM and Cer). These disturbances suggest a lasting metabolomic footprint beyond viral clearance, consistent with post-injury metabolic reprogramming (glycolytic shifts, lipid rewiring) that may continue to drive fibrogenesis ^22^.

A key finding of our study is the altered plasma amino acid profile in PWH with cirrhosis, persisting up to five years post-HCV eradication. We observed elevated levels of hydroxyproline, citrulline, and arginine, together with reduced levels of essential and branched-chain amino acids (e.g. L-valine, L-lysine, and L-tryptophan). Notably, increased arginine and citrulline, were already evident one year after treatment, suggesting early and sustained metabolic dysregulation despite viral clearance. These patterns align with reports linking arginine and citrulline levels to urea cycle and nitric oxide metabolism dysfunction in advanced liver disease ^23^. Impaired mitochondrial function disrupts citrulline-to-arginine conversion in the urea cycle, leading to citrulline accumulation and arginine diversion toward nitric oxide, which contributes to hyperammonemia, vascular dysregulation, hepatic encephalopathy and portal hypertension ^24, 25^. Reduced plasma levels of branched-chain amino acids have consistently been associated with liver dysfunction and hepatic encephalopathy ^24, 26, 27^. As branched-chain amino acids regulate gene expression, protein metabolism, apoptosis and hepatocyte regeneration, their depletion may signal impaired hepatocyte regenerative capacity in patients with advanced chronic liver disease ^28^. Lower circulating branched-chain amino acids may also reflect increased skeletal muscle uptake and mitochondrial impairment in hepatocytes ^29^. Collectively, these persistent amino acid alterations long after SVR suggest that viral clearance and time-alone do not resolve the underlying metabolic injury, suggesting ongoing subclinical liver damage or stalled metabolic recovery.

We found that PWH with cirrhosis showed sustained elevations in plasma levels of PC, PE, PS and PI, with little changes between one- and five-year post-treatment, while LPC, LPE and LPI levels remained largely unchanged. This suggest a stable pro-inflammatory glycerophospholipid profile associated with cirrhosis, consistent with prior links between LPC/LPE and hepatocyte lipotoxicity, pro-fibrogenic signaling, and sphingomyelin-ceramide axis alterations in fibrotic and cirrhotic states^30^. Although lipidomic studies in liver cirrhosis are limited, our data align with reports connecting glycerophospholipid dysregulation to liver disease and malignancies ^31–33^. For example, Clària et al. observed reduced conversion of PC to LPC in acute-on-chronic liver failure ^34^, and Cotte et al. reported increased PC and decreased LPC levels in cirrhotic patients with HCC. Specific changes we observed, such as increased PC 16:0/16:1 and lower LPC 20:4, have also been described in cirrhosis ^31^, while elevated PI plasma levels has been associated with cirrhosis by Meikle et al ^35^. Together, these findings support a persistent lipidomic signature in cirrhosis that may contribute to ongoing liver injury or systemic inflammation, highlighting potential targets for monitoring and therapy.

Although the overall glycerophospholipid profile remained stable, a key shift appeared five years post-therapy: increased desaturation of PC and PE species. Dysregulated desaturase activity is a hallmark of chronic liver disease, and altered phospholipid desaturation disrupts the balance between polyunsaturated and monounsaturated fatty acids in membrane phospholipids, a process known to affect lipid metabolism and contribute to aging and disease progression ^36^, promoting pro-inflammatory n-6 eicosanoid formation and increasing susceptibility to lipid peroxidation ^30^. This increased unsaturation of phospholipids sensitizes membranes to oxidative damage, a process well documented in advanced liver disease and decompensated cirrhosis ^37^. Oxidative lipid alterations have similarly been described across liver diseases, including decompensated cirrhosis ^37–39^. Thus, our findings suggest that these changes in glycerophospholipid composition are not markers of recovery but may reflect persistent metabolic disturbances that may drive liver disease progression long after HCV eradication.

A notable finding was the sustained reduction in TG species at both one and five years post-HCV therapy, undetected by routine biochemical tests that showed stable total plasma TG levels. This aligns with large-scale lipidomic studies reporting widespread TG species perturbations in cirrhosis^30^. As the liver plays a central role in TG synthesis, secretion, and very-low-density lipoprotein handling ^22, 40^, our findings support prior evidence of lower plasma TG levels in cirrhosis ^41^. Clinically, reduced TG levels has been associated with worse prognosis in HCC and advanced liver diseases ^30, 42^, underscoring its value as a prognostic marker. Thus, persistent TG depletion may reflect latent metabolic imbalance and ongoing hepatic dysfunction, offering potential as a sensitive long-term biomarker in liver disease management.

We also found stage-dependent alterations in sphingolipid metabolism, particularly in plasma levels of SM and Cer. Sphingolipids are bioactive lipids regulating membrane structure, inflammation and hepatic function ^43^. Prior lipidomic studies show SM increases in early phases or after interventions, while progressive fibrosis and cirrhosis are marked by Cer reductions and broader alterations in sphingolipid disturbances ^30^. Consistent with this, we observed a two-phase SM pattern: elevated at one year post-therapy but normalized by year five, as previously reported after HCV treatment and in advanced disease ^44^. In parallel, Cer levels declined progressively, aligning with evidence that lower ceramides correlate with fibrosis severity and increased HCC risk ^45^. The pronounced decrease in Cer at five years suggests persistent inflammation and ongoing liver disease progression despite viral clearance.

Our data also revealed a late increase in plasma acylcarnitine levels in PWH with cirrhosis, evident only five years post-HCV clearance, suggesting persistent mitochondrial dysfunction. Acylcarnitines are key to fatty acids β-oxidation, transporting fatty acids chains into mitochondria for energy production ^46, 47^. Normally, tightly regulated elevated medium- and long-chain acylcarnitines are a hallmark of impaired β-oxidation and oxidative stress, reported in decompensated cirrhosis and acute hepatic failure ^48–50^. Their progressive accumulation in our study population likely reflects ongoing mitochondrial and hepatic dysfunction long after HCV eradication.

## Conclusions

In conclusion, PWH with cirrhosis after HCV cure show a persistently altered metabolic profile, with most alterations remaining stable for up to five years. Newly emerging metabolic disturbances at this stage suggest ongoing liver disease progression despite HCV clearance. These findings underscore the need for long-term monitoring of this high-risk population, and support comprehensive metabolic profiling as a tool to identify individuals at risk and to better understand the persistent pathophysiology of liver disease after HCV clearance.

## Strengths and limitations

The main limitations are the modest sample size, which may have reduced the statistical power and patient retention issues that limited paired sampling. In addition, the cross-sectional design at each time point prevents causal inference. Despite these limitations, the study has notable strengths. Most importantly, it provides the first long-term metabolomic follow-up of PWH with cirrhosis after HCV cure, offering insights into their evolving metabolic landscape. This design enabled us to characterize a dynamic metabolic signature that shorter-term or less targeted studies would likely miss.

## Data Availability

The datasets used and analyzed during the current study may be made available by the corresponding author upon reasonable request.

## List of abbreviations

AIC: Akaike information criteria
AMR: Arithmetic mean ratio
CE-MS: Capillary Electrophoresis –mass spectrometry
Cer: Ceramides
CMM: CEU Mass Mediator
CV: Coefficient of variation
cART: Combination antiretroviral therapy
DAAs: Direct-acting antivirals
ESI: Electrospray ionization
FDR: False discovery rate
FA: Fatty acid
GLM: Generalized linear models
GL: Glycerolipids
GP: Glycerolphospolipids
HCV: Hepatitis C virus
HIV: Human immunodeficiency virus
IFN: Interferon
LMSD: LIPID MAPS Structure Database
LINEX: Lipid Network Explorer platform
LC-MS: Liquid chromatography–mass spectrometry
LSM: Liver stiffness measurement
LPC: Lysophosphatidylcholine
LPE: Lysophosphatidyletanolamine
LPI: Lysophosphatidylinositol
LPS: Lysophosphatidylserine
MeOH: Methanol
MTBE: Methyl-tert-butyl ether
OPLS-DA: Orthogonal partial least squares discriminant analysis
PWH: People with HIV
PA: Phosphatidic acid
PC: Phosphatidylcholine
PE: Phosphatidylethanolamines
PI: Phosphatidylinositol
PS: Phosphatidylserine
QC: Quality control
SP: Sphingolipids
SM: Sphingomyelin
SVR: Sustained virological response
TOF-MS: Time-of-flight Mass Spectrometer
TG: Triglycerides
UHPLC: Ultra-High-Performance Liquid-Chromatography
VIP: Variable importance in projection

## Declarations

### Ethics approval and consent to participate

This study was approved by the Research Ethics Committee of the Institute of Health Carlos III (CEI PI 72_2021-v2) and was carried out according to the Declaration of Helsinki.

### Consent for publication

Not applicable.

### Competing interests

The authors declare that they have no competing interests.

### Funding

This study was supported by grants from Instituto de Salud Carlos III (ISCIII; grant numbers CP17CIII/00007, PI18CIII/00028, PI21CIII/00033, and PI24CIII/00024 to MAJS, PI17/00657 and PI20/00474 to JB, PI17/00903 and PI20/00507 to JGG, and PI17CIII/00003 and PI20CIII/00004 to SR). The study was also funded by the CIBER -Consorcio Centro de Investigación Biomédica en Red- (CB 2021), Instituto de Salud Carlos III, Ministerio de Ciencia e Innovación and Unión Europea – NextGenerationEU (CB21/13/00044). R.M.-E. is César Nombela researcher supported and funded by Comunidad de Madrid (grant number 2023-T1/SAL-GL-28980).

## Acknowledgments

This study would not have been possible without the collaboration of all the patients, medical and nursing staff, and data managers who contributed to the project. We are especially grateful to the HIV BioBank, integrated into the Spanish AIDS Research Network and to all collaborating centers for the generous contribution of clinical samples to this work (see Appendix). All authors approved the final version of the article, including the authorship list.

During the preparation of this work the author(s) used Gemini 2.5 Pro in Google AI Studio and ChatGPT (OpenAI) in order to assist with language editing, grammar, and text flow. The conceptualization of the statistical analysis and the interpretation of the findings were performed entirely by the authors, who are fully responsible for the content of this paper.

## Authorship contribution

Funding body: MAJS and SR.

Study concept and design: MAJS.

Patients’ selection and clinical data acquisition: JB, JGG, CD, and VH.

Sample preparation, and biomarker analysis: AVB, DR, OBK, AFR, BR, CGR and CB.

Statistical analysis and interpretation of data: AVB, BR, CGR, RBL and MAJS.

Writing of the manuscript: AVB and MAJS.

Critical revision of the manuscript for relevant intellectual content: AFR, BR, CB, JB, RME and SR.

Supervision and visualization: MAJS and SR.

All authors approved the final version of the article.

## Authors’ information (optional)

Not applicable.

## Appendix

Members of the Marathon Study Group

**Hospital General Universitario Gregorio Marañón, Madrid:** A Carrero, P Miralles, JC López, F Parras, B Padilla, T Aldamiz-Echevarría, F Tejerina, C Díez, L Pérez-Latorre, C Fanciulli, I Gutiérrez, M Ramírez, S Carretero, JM Bellón, J Bermejo, and J Berenguer.

**Hospital Universitario La Paz, Madrid:** V Hontañón, JR Arribas, ML Montes, I Bernardino, JF Pascual, F Zamora, JM Peña, F Arnalich, M Díaz, J González-García.

**Hospital Universitari Vall d’Hebron, Barcelona:** E Van den Eynde, M Pérez, E Ribera, M Crespo.

**Hospital Universitario Príncipe de Asturias, Alcalá de Henares:** A Arranz, E Casas, J de Miguel, S Schroeder, J Sanz.

**Hospital Donostia, San Sebastián:** MJ Bustinduy, JA Iribarren, F Rodríguez-Arrondo, MA Von-Wichmann.

**Hospital Universitario de La Princesa, Madrid:** J Sanz, I Santos.

**Hospital Clínico San Carlos, Madrid:** J Vergas, MJ Téllez.

**Hospital Clínico Universitario, Valencia:** A Ferrer, MJ Galindo.

**Hospital Universitario Ramón y Cajal, Madrid:** JL Casado, F Dronda, A Moreno, MJ Pérez-Elías, MA Sanfrutos, S Moreno, C Quereda.

**Hospital General Universitario, Valencia:** L Ortiz, E Ortega.

**Fundación SEIMC-GESIDA, Madrid:** M Yllescas, P Crespo, E Aznar, H Esteban.

## Notes

### Competing Interest Statement

The authors have declared no competing interest.

### Funding Statement

This study was funded by grants from Instituto de Salud Carlos III (ISCIII; grant numbers CP17CIII/00007, PI18CIII/00028, PI21CIII/00033, and PI24CIII/00024 to MAJS, PI17/00657 and PI20/00474 to JB, PI17/00903 and PI20/00507 to JGG, and PI17CIII/00003 and PI20CIII/00004 to SR). The study was also funded by the CIBER Consorcio Centro de Investigacion Biomedica en Red (CB 2021), Instituto de Salud Carlos III, Ministerio de Ciencia e Innovacion and Union Europea NextGenerationEU (CB21/13/00044). R.M.-E. is Cesar Nombela researcher supported and funded by Comunidad de Madrid (grant number 2023-T1/SAL-GL-28980).

## References

1. Liu CH, Kao JH. Acute hepatitis C virus infection: clinical update and remaining challenges. Clin Mol Hepatol. Jul 2023;29(3):623–642.

2. Rossi C, Jeong D, Wong S, et al. Sustained virological response from interferon-based hepatitis C regimens is associated with reduced risk of extrahepatic manifestations. J Hepatol. Dec 2019;71(6):1116–1125.

3. Mazzaro C, Quartuccio L, Adinolfi LE, et al. A Review on Extrahepatic Manifestations of Chronic Hepatitis C Virus Infection and the Impact of Direct-Acting Antiviral Therapy. Viruses. Nov 9 2021;13(11).

4. Lockart I, Yeo MGH, Hajarizadeh B, Dore GJ, Danta M. HCC incidence after hepatitis C cure among patients with advanced fibrosis or cirrhosis: A meta-analysis. Hepatology. Jul 2022;76(1):139–154.

5. Chang B, Tian H, Huang A, et al. Prevalence and prediction of hepatocellular carcinoma in alcohol-associated liver disease: a retrospective study of 136 571 patients with chronic liver diseases. eGastroenterology. Jan 2024;2(1):e100036.

6. Chen YT, Chen TI, Yang TH, et al. Long-term Risks of Cirrhosis and Hepatocellular Carcinoma Across Steatotic Liver Disease Subtypes. Am J Gastroenterol. Nov 1 2024;119(11):2241–2250.

7. Shen C, Jiang X, Li M, Luo Y. Hepatitis Virus and Hepatocellular Carcinoma: Recent Advances. Cancers (Basel). Jan 15 2023;15(2).

8. Verna EC, Morelli G, Terrault NA, et al. DAA therapy and long-term hepatic function in advanced/decompensated cirrhosis: Real-world experience from HCV-TARGET cohort. J Hepatol. Sep 2020;73(3):540–548.

9. Cambiaghi A, Ferrario M, Masseroli M. Analysis of metabolomic data: tools, current strategies and future challenges for omics data integration. Brief Bioinform. May 1 2017;18(3):498–510.

10. Wang R, Li B, Lam SM, Shui G. Integration of lipidomics and metabolomics for in-depth understanding of cellular mechanism and disease progression. J Genet Genomics. Feb 20 2020;47(2):69–83.

11. Darweesh M, Mohammadi S, Rahmati M, Al-Hamadani M, Al-Harrasi A. Metabolic reprogramming in viral infections: the interplay of glucose metabolism and immune responses. Front Immunol. 2025;16:1578202.

12. Deme P, Rubin LH, Yu D, et al. Immunometabolic Reprogramming in Response to HIV Infection Is Not Fully Normalized by Suppressive Antiretroviral Therapy. Viruses. Jun 15 2022;14(6).

13. Mu W, Patankar V, Kitchen S, Zhen A. Examining Chronic Inflammation, Immune Metabolism, and T Cell Dysfunction in HIV Infection. Viruses. Jan 31 2024;16(2).

14. Medrano LM, Garcia-Broncano P, Berenguer J, et al. Elevated liver stiffness is linked to increased biomarkers of inflammation and immune activation in HIV/hepatitis C virus-coinfected patients. AIDS. Jun 1 2018;32(9):1095–1105.

15. Castera L, Forns X, Alberti A. Non-invasive evaluation of liver fibrosis using transient elastography. J Hepatol. May 2008;48(5):835–847.

16. Gonzalez-Riano C, Gradillas A, Barbas C. Exploiting the formation of adducts in mobile phases with ammonium fluoride for the enhancement of annotation in liquid chromatography-high resolution mass spectrometry based lipidomics. Journal of Chromatography Open. 2021;1:100018.

17. Mamani-Huanca M, de la Fuente AG, Otero A, et al. Enhancing confidence of metabolite annotation in Capillary Electrophoresis-Mass Spectrometry untargeted metabolomics with relative migration time and in-source fragmentation. J Chromatogr A. Jan 4 2021;1635:461758.

18. Gil-de-la-Fuente A, Godzien J, Saugar S, et al. CEU Mass Mediator 3.0: A Metabolite Annotation Tool. J Proteome Res. Feb 1 2019;18(2):797–802.

19. Fernandez Requena B, Nadeem S, Reddy VP, et al. LiLA: lipid lung-based ATLAS built through a comprehensive workflow designed for an accurate lipid annotation. Commun Biol. Jan 5 2024;7(1):45.

20. Kuligowski J, Sanchez-Illana A, Sanjuan-Herraez D, Vento M, Quintas G. Intra-batch effect correction in liquid chromatography-mass spectrometry using quality control samples and support vector regression (QC-SVRC). Analyst. Nov 21 2015;140(22):7810–7817.

21. Kohler N, Rose TD, Falk L, Pauling JK. Investigating Global Lipidome Alterations with the Lipid Network Explorer. Metabolites. Jul 28 2021;11(8).

22. Horn CL, Morales AL, Savard C, Farrell GC, Ioannou GN. Role of Cholesterol-Associated Steatohepatitis in the Development of NASH. Hepatol Commun. Jan 2022;6(1):12–35.

23. Zhang Y, Higgins CB, Tica S, et al. Hierarchical tricarboxylic acid cycle regulation by hepatocyte arginase 2 links the urea cycle to oxidative metabolism. Cell Metab. Sep 3 2024;36(9):2069–2085 e2068.

24. Espina S, Gonzalez-Irazabal Y, Sanz-Paris A, et al. Amino Acid Profile in Malnourished Patients with Liver Cirrhosis and Its Modification with Oral Nutritional Supplements: Implications on Minimal Hepatic Encephalopathy. Nutrients. Oct 25 2021;13(11).

25. Tang J, Xiong K, Zhang T, Han H. Application of Metabolomics in Diagnosis and Treatment of Chronic Liver Diseases. Crit Rev Anal Chem. 2022;52(5):906–916.

26. Wu T, Wang M, Ning F, et al. Emerging role for branched-chain amino acids metabolism in fibrosis. Pharmacol Res. Jan 2023;187:106604.

27. Mansoori S, Ho MY, Ng KK, Cheng KK. Branched-chain amino acid metabolism: Pathophysiological mechanism and therapeutic intervention in metabolic diseases. Obes Rev. Feb 2025;26(2):e13856.

28. Tajiri K, Shimizu Y. Branched-chain amino acids in liver diseases. World J Gastroenterol. Nov 21 2013;19(43):7620–7629.

29. Zhang Y, Zhan L, Zhang L, Shi Q, Li L. Branched-Chain Amino Acids in Liver Diseases: Complexity and Controversy. Nutrients. Jun 14 2024;16(12).

30. Musso G, Saba F, Cassader M, Gambino R. Lipidomics in pathogenesis, progression and treatment of nonalcoholic steatohepatitis (NASH): Recent advances. Prog Lipid Res. Jul 2023;91:101238.

31. Cotte AK, Cottet V, Aires V, et al. Phospholipid profiles and hepatocellular carcinoma risk and prognosis in cirrhotic patients. Oncotarget. Mar 15 2019;10(22):2161–2172.

32. Kartsoli S, Kostara CE, Tsimihodimos V, Bairaktari ET, Christodoulou DK. Lipidomics in non-alcoholic fatty liver disease. World J Hepatol. Aug 27 2020;12(8):436–450.

33. Babu AF, Palomurto S, Karja V, et al. Metabolic signatures of metabolic dysfunction-associated steatotic liver disease in severely obese patients. Dig Liver Dis. Dec 2024;56(12):2103–2110.

34. Claria J, Curto A, Moreau R, et al. Untargeted lipidomics uncovers lipid signatures that distinguish severe from moderate forms of acutely decompensated cirrhosis. J Hepatol. Nov 2021;75(5):1116–1127.

35. Meikle PJ, Mundra PA, Wong G, et al. Circulating Lipids Are Associated with Alcoholic Liver Cirrhosis and Represent Potential Biomarkers for Risk Assessment. PLoS One. 2015;10(6):e0130346.

36. Choudhary RC, Kuschner CE, Kazmi J, et al. The Role of Phospholipid Alterations in Mitochondrial and Brain Dysfunction after Cardiac Arrest. Int J Mol Sci. Apr 24 2024;25(9).

37. Engelmann C, Claria J, Szabo G, Bosch J, Bernardi M. Pathophysiology of decompensated cirrhosis: Portal hypertension, circulatory dysfunction, inflammation, metabolism and mitochondrial dysfunction. J Hepatol. Jul 2021;75 Suppl 1(Suppl 1):S49-S66.

38. de Andrade KQ, Moura FA, dos Santos JM, de Araujo OR, de Farias Santos JC, Goulart MO. Oxidative Stress and Inflammation in Hepatic Diseases: Therapeutic Possibilities of N-Acetylcysteine. Int J Mol Sci. Dec 18 2015;16(12):30269–30308.

39. Pomacu MM, Trasca MD, Padureanu V, et al. Interrelation of inflammation and oxidative stress in liver cirrhosis. Exp Ther Med. Jun 2021;21(6):602.

40. Alves-Bezerra M, Cohen DE. Triglyceride Metabolism in the Liver. Compr Physiol. Dec 12 2017;8(1):1–8.

41. Wang P, Wang Y, Liu H, et al. Role of triglycerides as a predictor of autoimmune hepatitis with cirrhosis. Lipids Health Dis. Oct 25 2022;21(1):108.

42. Liu X, Li M, Wang X, et al. Effect of serum triglyceride level on the prognosis of patients with hepatocellular carcinoma in the absence of cirrhosis. Lipids Health Dis. Nov 6 2018;17(1):248.

43. Ishay Y, Nachman D, Khoury T, Ilan Y. The role of the sphingolipid pathway in liver fibrosis: an emerging new potential target for novel therapies. Am J Physiol Cell Physiol. Jun 1 2020;318(6):C1055–C1064.

44. Nojima H, Shimizu H, Murakami T, Shuto K, Koda K. Critical Roles of the Sphingolipid Metabolic Pathway in Liver Regeneration, Hepatocellular Carcinoma Progression and Therapy. Cancers (Basel). Feb 20 2024;16(5).

45. Horing M, Peschel G, Grimm J, et al. Serum Ceramide Species Are Associated with Liver Cirrhosis and Viral Genotype in Patients with Hepatitis C Infection. Int J Mol Sci. Aug 29 2022;23(17).

46. Zhou L, Wang Q, Yin P, et al. Serum metabolomics reveals the deregulation of fatty acids metabolism in hepatocellular carcinoma and chronic liver diseases. Anal Bioanal Chem. Apr 2012;403(1):203–213.

47. Dambrova M, Makrecka-Kuka M, Kuka J, et al. Acylcarnitines: Nomenclature, Biomarkers, Therapeutic Potential, Drug Targets, and Clinical Trials. Pharmacol Rev. Jul 2022;74(3):506–551.

48. Miyaaki H, Kobayashi H, Miuma S, et al. Blood carnitine profiling on tandem mass spectrometry in liver cirrhotic patients. BMC Gastroenterol. Feb 19 2020;20(1):41.

49. Zhang IW, Sanchez-Rodriguez MB, Lopez-Vicario C, et al. Palmitoylcarnitine impairs immunity in decompensated cirrhosis. JHEP Rep. Nov 2024;6(11):101187.

50. Gao B, Argemi J, Bataller R, Schnabl B. Serum Acylcarnitines Associated with High Short-Term Mortality in Patients with Alcoholic Hepatitis. Biomolecules. Feb 14 2021;11(2).

