## Supplementary Data for "Distinct metabolomic and lipidomic profiles associated with cirrhosis after HCV cure in people with HIV: findings at one and five years"

**Supplementary Data 1.** Extension of methods section.

#### *1. Reagents and standards*

LC-MS grade methanol (MeOH), acetonitrile (ACN), and isopropanol (IPA) were obtained from Fisher Scientific (Pennsylvania, United States). Analytical grade ammonia solution (28%, GPR RECTAPUR®) and acetic acid glacial (AnalaR® NORMAPUR®) were obtained from VWR Chemicals (Pennsylvania, United States). The ammonium fluoride (NH4F) (ACS reagent, ≥ 98%) and Methyl-tert-butyl ether (MTBE) were purchased from Sigma-Aldrich (Steinheim, Germany). Reverse-osmosed ultrapure water for aqueous solutions was obtained from a Milli-Qplus185 system (Millipore, Billerica, MA, USA). For analytical quality assurance, C17–Sphinganine and $d_{31}$–palmitic acid were purchased from Avanti Polar Lipids (Alabaster, Alabama, USA) for LC-MS. In the same way, for CE-MS quality assurance, methionine sulfone, paracetamol, 2-(N-morpholino)ethanesulfonic acid (MES), and formic acid were acquired from Sigma (Steinheim, Germany). Reference mass solutions for LC-MS and CE-MS were obtained from Agilent Technologies.

#### *2. Blood samples*

Peripheral blood samples were collected in EDTA tubes at two points: one and five years after successful completion of HCV therapy. Plasma samples were separated by Ficoll-Paque density gradient and stored at -80ºC in the Spanish HIV HGM Biobank until shipment for its use.

#### *3. Viral inactivation*

Viral inactivation of plasma samples was performed by mixing 300 µL of plasma with 900 µL of cold MeOH:EtOH (1:1, v/v), reaching a 1:3, v/v ratio. After that, the samples were vortex mixed for 1 min and introduced on an ice-bath for 5 min. Finally, the samples were centrifuged at 16000g, during 20 min and 4ºC, and then they were stored at -80ºC until analysis.

#### *4. Lipidomics Analysis*

##### *4.1. Lipid extraction*

Inactivated plasma samples were prepared at the "Centro de Metabolómica y Bioanálisis, CEMBIO" (Madrid, Spain), following an extraction method consisting of MeOH:MTBE (1:1, v/v), which was tested and developed in our laboratory. Briefly, the samples were thawed on ice and vortex-mixed for 2 minutes. On the ice, we pipetted 200 µL of plasma sample, consisting of 50 µL of plasma and 150 µL of MeOH:EtOH (1:1, v/v), to an Eppendorf tube. Then, we added 175 µL of MTBE (RT) and vortex-mixed samples for 30 min. In addition, a mixture of nonendogenous internal standards (1 ppm of C17-Sphinganine and 2 ppm d31-palmitic acid) was added to each sample. Following, the samples were centrifuged for 15 minutes at

16,1 rpm and 15ºC. We transferred 100 µL of the supernatant into clean HPLC-MS (Thermo Fisher Scientific, Madrid, Spain) vials with glass insert and centrifuged at 2000g and 15ºC for 5 min before injecting into the system.

#### *4.2.* Quality Management Assurance and Blank samples

Quality Control (QC) samples were prepared by polling equal volumes of each plasma sample, 30 µL in our case, to an Eppendorf tube. These QC samples were then processed in parallel with the rest of the experimental samples in the same manner. During the run, the system's stability, performance, and sample treatment method repeatability were all monitored. Along with the other samples, two blank samples were prepared using the same lipid extraction procedure with the sample solvents and were injected at the beginning and end of the analytical sequence to find common contaminations.

#### *4.3. Analytical conditions selected for a Lipidomics Analysis.*

We performed an untargeted lipidomics analysis to cover the broader spectrum of plasma lipidome. Samples were analyzed using an Agilent 1290 Infinity II Ultra-High-Performance Liquid-Chromatography (UHPLC) system coupled to an Agilent 6546 Quadrupole Time-of-Flight (QTOF) Mass Spectrometer (MS) equipped with dual Agilent Jet Stream (AJS) Electrospray (ESI) ion source. The Agilent 1290 Infinity II Multisampler system was used to uptake 1 µL in ESI(+) and ESI(-) of the extracted plasma samples, set at a temperature of 15 °C to avoid lipid precipitation. An Agilent InfinityLab Poroshell 120 EC – C18 (3.0 ×100 mm, 2.7 µm) (Agilent Technologies) reverse phase column and compatible guard column (Agilent InfinityLab Poroshell 120 EC –C18, 3.0 ×5 mm, 2.7 µm) was used and held at 50 °C(1).

The Agilent 6546 QTOF mass spectrometer, equipped with a dual AJS ESI ion source, setting parameters were as follows: 150 V fragmentor, 65 V skimmer, 3500 V capillary voltage, 750 V octopole radio frequency voltage, 10 L/min nebulizer gas flow, 200 °C gas temperature, 50 psi nebulizer gas pressure, 12 L/min sheath gas flow, and 300 °C sheath gas temperature. Data were collected in separate runs in positive and negative ESI modes, operated in full scan mode from 40 to 1700 *m/z* with a scan rate of 3 spectra/s. A solution consisting of two reference mass compounds was used throughout the whole analysis: purine ($C_5H_4N_4$) at *m/z* 121.0509 for ESI(+) and *m/z* 119.0363 for ESI(-); and HP-0921 ($C_{18}H_{18}O_6N_3P_3F_{24}$) at *m/z* 922.0098 for ESI(+) and *m/z* 980.0163 (HP-0921 + acetate) for ESI(-). An Agilent 1260 Iso Pump was used to continuously infuse the masses into the system at a 1 mL/min (split ratio 1:100) to provide a constant mass correction(1).

The mobile phases used for both positive and negative ionization modes consisted of (A) 10 mM ammonium acetate and 0.2 mM $NH_4F$ in $H_2O$/MeOH (9:1, v/v) and (B) 10 mM ammonium acetate, 0.2 mM $NH_4F$ in ACN/MeOH/IPA (2:3:5, v/v/v). A solvent mixture of MeOH:IPA (50:50, v/v) was used for the multiwash strategy, with a wash time set at 15 s and an A/B (30:70, v/v) mixture to assist in the starting conditions. The chromatography gradient started at 70% of B at 0 –1 min, 86% B at 3.5 –10 min, and 100% B at 11–17 min. Starting conditions were recovered by min 17, followed by a 2 min re-equilibration time, reaching a total running time of 19 min. The flow rate was kept constant at 0.6 mL/min during the analysis.

At the end of the analysis, iterative MS/MS acquisition mode was performed for both ionization modes using QC samples. Two different collision energies, 20 eV and 40 eV, were used to conduct ten measurements (five measurements at each one), and they were operated with an MS and MS/MS scan rate of 3 spectra/s, 3 precursors per cycle, a mass range of m/z 40 - 1700, a narrow (~ 1.3 amu) MS/MS isolation width, and 5000 counts and 0.001 % of MS/MS threshold. The software chooses the three more intense precursor ions for subsequent fragmentation to obtain their MS/MS spectra in each measurement. In the second measurement of the same sample, the software discards the first three previously selected precursor ions and selects the next three more intense precursor ions at the same precise time point. In addition, reference masses and contaminants detected in blank samples were excluded, preventing their inclusion in the iterative MS/MS runs. By analyzing the sample many times, we were able to obtain the MS/MS information of most of the plasma lipidome by collecting thousands of MS/MS spectra using MassHunter Workstation Software LC-MS Data Acquisition v B.09.00 (Agilent Technologies, Waldrobonn, Germany).

#### *4.4. Lipid Annotation process*

The lipid Annotation process was performed following a lipid annotation strategy developed by our laboratory (2). This strategy consisted of the use of a combination of bioinformatics tools based on three different annotation mechanisms: spectra matching (Lipid Annotator, MS-Dial), the bottom-up strategy (LipidHunter) and the fragment intensity rules (LipidMS). Following this approach, we significantly improved the lipid annotation process. The software parameters were set as follows:

- *Lipid Annotator v1.0* (Agilent) (3): Q-Score ≥ 20.0, adduct selection $H^+$, $Na^+$ and $NH_4^+$ for positive ion mode and $H^-$ and $C_2H_3O_2^-$ for negative ion mode. We selected all lipid classes to perform an untargeted analysis. For the ID parameters, the Mass Threshold was set at mass deviation ≤ 20.0 ppm, the "Report top candidate only" option was selected, the Fragment score was ≥ 30, the Total score was ≥ 60, and the Constituent Level was ≥ 10%.
- *MS-DIAL 4* (Riken) (4): the raw data files were converted into "*.ibf*" files using the MS IBF file Converter software. For MS/MS data reprocessing, we selected the soft ionization for LC-MS/MS, chromatography separation type, conventional LC-MS method type, and profile data as the data type. We selected the corresponding ionization mode (positive or negative) for each analysis and, Lipidomics was used as the target omics. The MS1 and MS/MS *m/z* detection window was set at 40 – 1700 Da, and

the retention time window was set at 0 – 19 minutes. The smoothing level for the peak detection window was set as 1 scan. The Accurate mass tolerance was 0.01 Da for MS1 and 0.025 for MS2 for the Identification window, and the identification score cut-off was set at 70%. Next, we selected specific adducts depending on the ionization mode we analyzed ($H^+$, $Na^+$ and $NH_4^+$ for positive and $H^-$ and $C_2H_3O_2^-$ for negative ion mode). The rest of the parameters were set as default.

- *LipidHunter* (5): we set from 0 – 19 min the scan range, 40 – 1700 *m/z* range, +/- 0.75 *m/z* precursor window, DDA Top 6, +/- 20 ppm for MS tolerance level, the absolute intensity for the MS level threshold was set at 1000, +/- 20 ppm MS/MS tolerance level, the absolute intensity for the MS/MS level threshold was set at 10, 80% isotope score, 75% Rank score and 0.10% as the minimum relative intensity for the scoring.
- *LipidMS 3.0* (6, 7)*:* We reprocessed our data by selecting the Batch processing option in the corresponding ionization mode. The *m/z* tolerance for MS1 and MS/MS was set at 20 ppm. The tolerance for the RT window was set at 30 seconds. The rest of the parameters were set as default. Finally, the lipid classes selected for the annotation process were established according to the ionization mode.

Subsequently, redundant data and false positive annotations were eliminated by combining and manually curating the collected information. In addition, the tentative annotation provided by the software annotation tools was combined with the manual inspection of the MS and MS/MS spectra data of the samples, based on the fundaments of structural elucidation and the assistance of the CEU Mass Mediator (CMM) free access technology (8) to corroborate the accuracy of the lipid annotations.

### 5. Capillary Electrophoresis Analysis

#### 5.1. Metabolites extraction

Inactivated plasma samples were also prepared at the "Centro de Metabolómica y Bioanálisis, CEMBIO" (Madrid, Spain). The samples were thawed on ice and vortex-mixed for 2 min. The samples were centrifuged for 15 min at 4ºC and 16,000xg, and 400 µL of the supernatant was transferred to an Eppendorf tube. Next, the samples were dried on the SpeedVac Concentrator (Thermo Fisher Scientific, Waltham, MA, USA) and resuspended in 200 µL of 0.1M Formic acid containing 0.2 mM methionine sulfone, 1 mM paracetamol, 0.25 mM (MES) and 25% ACN as internal standards (IS). Samples were then vortex-mixed for 1 min and transferred to a 30-kDa protein cut-off filter for the deproteinization process through a centrifugation step using a Centrifree ultracentrifugation device (Millipore Ireland Ltd., Cork, Ireland) for 90 min at 2000xg and 4ºC. The filtrate was transferred to a chromacol vial for CE-MS analysis. The vials were then centrifuged for 10 min at 2000xg and 4ºC.

#### *5.2.* Quality Management Assurance and Blank samples

Equal volumes of each plasma sample (50 µL) were pooled into an Eppendorf tube to create Quality Control (QC) samples. QC samples were then prepared and processed in parallel with the rest of the plasma samples, following the metabolites extraction protocol described above. The system's stability, performance, and sample treatment method repeatability were all monitored during the run. Additionally, two blank samples were prepared along with the other samples using the same metabolite extraction procedure with the sample solvents. As was explained before, the blank samples were analyzed at the beginning and end of the analytical sequence to find potential contaminations.

#### ***5.3. Analytical conditions selected for a Metabolomics Analysis.***

An untargeted metabolomics-based approach was employed to comprehensively capture the diverse spectrum of plasma metabolites. Samples were analyzed using an Agilent CE 7100 coupled with an Agilent 6224 time-of-flight Mass Spectrometer (TOF-MS) analyzer. The coupling was performed with an electrospray source, helped by a sheath liquid, which consisted of methanol/water (1/1, v/v), formic acid (1 mM), and two reference masses (purine, m/z 121.050873; HP-0921, m/z 922.009798). The auxiliary sheath liquid, supplied by an Agilent 1200 ISO Pump, was used to compensate the volume and increase the necessary volatility for MS. For CE-MS separation analysis in positive ionization mode, a fused-silica capillary (Agilent Technologies; total length 100 cm x 50 µm i.d. x 360 µm) was used. First of all, a background electrolyte (BGE) (1 M formic acid in 10% methanol solution) solution was flushed for 5 min (950 mbar) through the capillary to condition and rinse it. A voltage of 30 kV was applied to displace the BGE ions during 10 s. Samples were then injected over 50s at 50mbar, followed by the BGE injection for 10 s at 100 mbar to improve the reproducibility of the analysis. The separation was carried out at a pressure of 25 mbar and a voltage of +30kV, in positive ionization mode, with an observed current of 24 µA under these conditions.

When the metabolites leave the capillary, a positively charger spray is formed with a flow rate of 0.6 mL/min (1:100) of the auxiliary liquid at 10 spig nebulization pressure with nitrogen and 3500 V of capillary voltage. The spray was dried with a hot nitrogen flow of 10 mL/min at 200 °C. The ions formed were directed to the TOF by 125 V - for adducts, dimmers and transformation data - and 200 V - for the acquisition of in-source fragment ions - fragmentor voltages, skimmer (65 V) and octopole (750 V). The MS was operated in ESI (+) positive polarity mode and a full range mass from 50 and 1050 Da was acquired at a scanning rate of 1.02 scans/s. Finally, selected samples were analyzed at the end of the analytical run by applying a higher fragmentor voltage (125,150,175 and 200 V) to acquire in-source fragment ions (pseudo-MS/MS). By adopting this approach, the obtained spectrum is similar to a 10 eV MS/MS spectrum, although some fragment ions could differ(9, 10). The total time of the analytical run was 35 min. The CE-

MS system was controlled by the MassHunter Workstation 6200 series TOF version B.09.00 c(B9044.0) acquisition software.

#### *5.4. Metabolite Annotation process*

Metabolite annotations were conducted utilizing a predefined CE-MS database meticulously tailored for CE-MS (9). This comprehensive in-house database is integrated into the CEU Mass Mediator (CMM), an open-access resource, and was imported into the Agilent MassHunter Profinder software (B.10.0.2, Agilent Technologies, Santa Clara, CA, USA) for CE-MS data reprocessing. Metabolites were reported in agreement with the Metabolomics Standards Initiative (MSI) criteria (11).

### *6.* Data Reprocessing

The raw data files were reprocessed by performing the hybrid strategy (2), which consisted of using an extensive compound database, both lipidomics and metabolomics data, respectively, which was imported into the Agilent MassHunter Profinder software (B.10.0.2, Agilent Technologies, Santa Clara, CA, USA) to perform the time alignment and feature extraction using the "Batch Targeted Feature Extraction" mode. This strategy allowed us to ascertain the differences in the plasma lipidomic and metabolomic fingerprinting. The migration time (MT), monoisotopic exact mass, and molecular formula were all included in the database. The clustering of coeluting ions connected by charge-state, isotopic distribution, and/or the presence of various adducts and dimers in the analyzed samples was used for the feature-building process. The following adducts were selected for lipidomics analysis to detect coeluting adducts of the same feature: [M+H]+, [M+Na]+, [M+K]+, [M+NH4]+ and [M+C2H6N2+H]+ for positive ionization mode; [M-H]-, [M+Cl]-, [M+CH3COOH-H]-, and [M+CH3COONa-H]− for negative ionization. In the case of positive CE-MS ionization mode, [M+H]+ and M+Na]+ were selected. Common organic molecules (no halogens) for LC-ESI(+)-MS and CE-ESI(+)-MS and common organic molecules for LC-ESI (-)-MS were used as the isotope grouping pattern, and the charge state was restricted to 1 - 2. We established the masses match tolerance at ± 20 ppm and ± 0.5 min for the RT. The RT score accounted for 100.00% of the total score. To complete the integration, we selected the "Agile 2" algorithm. The data format chromatogram employed was the centroid. The peak spectrum parameters were as follows: average scans > 10% of peak height, TOF spectra were excluded if above 20.0 % of saturation in the m/z ranges used, and empty spectrum will never return. The mass spectral data type was centroid. As a final post-processing filter, we only considered the features present in at least 50% of the samples within a single sample group.

#### *6.1. Data normalization and filtering*

Regarding lipidomics data, Data normalization was conducted utilizing the Kuligowski transformation (12) implemented in MATLAB (R2023a, MathWorks). This chemometric technique corrects batch effects post-acquisition, mainly targeting variability in QC samples and mitigating observed trends. For CE-MS metabolite analysis, normalization was achieved using the methionine sulfone IS, added during the initial sample preparation phase. Subsequently, lipid species and metabolites were selected based on their coefficient of variation (CV) within the QC samples, applying a threshold of 30%.

#### *6.2. Statistical analysis*

Statistical analysis was performed using MetaboAnalyst 6.0 software (http://www.metaboanalyst.ca/) and R software version 4.3.1 (R Foundation for Statistical Computing, Vienna, Austria).

For descriptive analysis, categorical variables were expressed as absolute count (percentage), and continuous variables were shown as median (interquartile range). Chi-square and Mann-Whitney U tests were used to compare categorical and quantitative variables, respectively.

For the multivariate analysis, firstly, metabolomic variables were log-transformed (log2) and auto-scaled. Next, we performed a supervised multivariate analysis by orthogonal partial least squares discriminant analysis (OPLS-DA), which models simultaneously all features. We obtained variable importance in projection (VIP) score for each feature and model validity was assessed using 1000 permutation test.

For the association analysis, Generalized Linear Models (GLM) with gamma distribution was performed to evaluate the association between metabolomic features (dependent variable) and cirrhosis (LSM≥12.5 KPa) (independent variable) one year and five years after successful completion of HCV therapy. A forward stepwise method, based on the lowest Akaike information criteria (AIC), was used to adjust for relevant covariates, including age, gender, BMI, and HCV treatment. Results were reported as arithmetic means ratio (AMR) and their corresponding levels of significance. Multiple testing correction was performed using the false discovery rate (FDR) according to Benjamini and Hochberg procedure. Statistical significance was defined as $p<0.05$ and q-value<0.2.

REFERENCES


1. Gonzalez-Riano C, Gradillas A, and Barbas C. Exploiting the Formation of Adducts in Mobile Phases with Ammonium Fluoride for the Enhancement of Annotation in Liquid Chromatography High-Resolution Mass Spectrometry (LCHR-MS)-based Lipidomics. *Journal of Chromatography Open.* 2021:100018.
2. Fernández Requena B, Nadeem S, Reddy VP, Naidoo V, Glasgow JN, Steyn AJ, et al. LiLA: lipid lung-based ATLAS built through a comprehensive workflow designed for an accurate lipid annotation. *Communications Biology.* 2024;7(1):45.
3. Koelmel JP, Li X, Stow SM, Sartain MJ, Murali A, Kemperman R, et al. Lipid annotator: towards accurate annotation in non-targeted liquid chromatography high-resolution tandem mass spectrometry (LC-HRMS/MS) lipidomics using a rapid and user-friendly software. *Metabolites.* 2020;10(3):101.
4. Tsugawa H, Ikeda K, Takahashi M, Satoh A, Mori Y, Uchino H, et al. MS-DIAL 4: accelerating lipidomics using an MS/MS, CCS, and retention time atlas. *BioRxiv.* 2020:2020.02. 11.944900.
5. Ni Z, Angelidou G, Lange M, Hoffmann R, and Fedorova M. LipidHunter identifies phospholipids by high-throughput processing of LC-MS and shotgun lipidomics datasets. *Analytical Chemistry.* 2017;89(17):8800-7.
6. Alcoriza-Balaguer MI, García-Cañaveras JC, López An, Conde I, Juan O, Carretero Jn, et al. LipidMS: an R package for lipid annotation in untargeted liquid chromatography-data independent acquisition-mass spectrometry lipidomics. *Analytical chemistry.* 2018;91(1):836-45.
7. Alcoriza-Balaguer MI, García-Cañaveras JC, Ripoll-Esteve FJ, Roca M, and Lahoz A. LipidMS 3.0: an R-package and a web-based tool for LC-MS/MS data processing and lipid annotation. *Bioinformatics.* 2022;38(20):4826-8.
8. Gil-De-La-Fuente A, Godzien J, Saugar S, Garcia-Carmona R, Badran H, Wishart DS, et al. CEU Mass Mediator 3.0: A Metabolite Annotation Tool. *Journal of Proteome Research.* 2019;18(2):797-802.
9. Mamani-Huanca M, de la Fuente AG, Otero A, Gradillas A, Godzien J, Barbas C, et al. Enhancing confidence of metabolite annotation in capillary electrophoresis-mass spectrometry untargeted metabolomics with relative migration time and in-source fragmentation. *Journal of Chromatography A.* 2021;1635:461758.
10. Mamani-Huanca M, Gradillas A, Gil de la Fuente A, López-Gonzálvez An, and Barbas C. Unveiling the fragmentation mechanisms of modified amino acids as the key for their targeted identification. *Analytical chemistry.* 2020;92(7):4848-57.
11. Sumner LW, Amberg A, Barrett D, Beale MH, Beger R, Daykin CA, et al. Proposed minimum reporting standards for chemical analysis Chemical Analysis Working Group (CAWG) Metabolomics Standards Initiative (MSI). *Metabolomics.* 2007;3(3):211-21.
12. Kuligowski J, Sanchez-Illana A, Sanjuan-Herraez D, Vento M, and Quintas G. Intra-batch effect correction in liquid chromatography-mass spectrometry using quality control samples and support vector regression (QC-SVRC). *Analyst.* 2015;140(22):7810-7.

**Supplementary Data 2. A)** Evolution of LSM values from baseline (beginning of HCV therapy) to five years after completion of successful HCV treatment in patients with HIV (PWH) stratified by cirrhosis status (LSM≥12.5kPa). **Statistics**: p-value was calculated by the Wilcoxon test and Mann Whitney tests. **B)** Individual evolution of LSM values from one year to five years after HCV therapy in PWH. **C)** PWH with cirrhosis at one and five years after completion HCV therapy. **Abbreviations:** LSM, liver stiffness measurement.

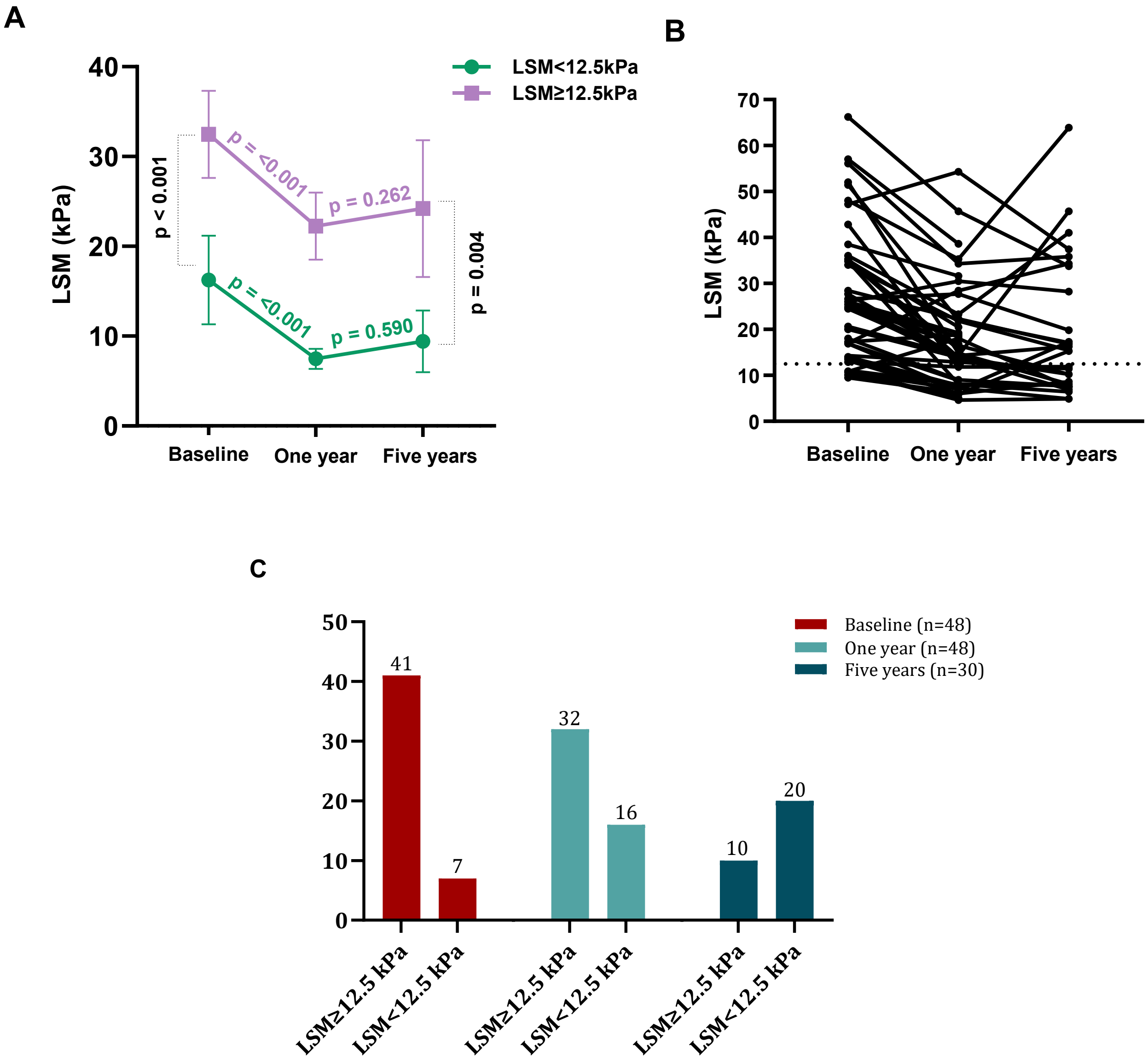

**Supplementary Data 3**. Biochemical clinical data of people living with HIV (PLW) at one year after completion of HCV therapy categorized by cirrhosis (LSM≥12.5kPa). **Statistics**: The values are expressed as the absolute number (percentage) and median (interquartile range). **Abbreviations**: MCV, mean corpuscular volume; INR, international normalized ratio; AST, aspartate transaminase; ALT, alanine aminotransferase; HDL, high-density lipoprotein; LDL, low-density lipoprotein.

| | **All patients** | **LSM<12.5kPa** | **LSM≥12.5kPa** | **p-value** |
|---|---|---|---|---|
| **No.** | 48 | 16 | 32 | |
| Neutrophils | 2640 (2250-3515) | 2850 (2430-3275) | 2600 (2105-3550) | 0.369 |
| Platelets ($10^3$) | 122 (93-163.5) | 141.5 (125.8-198) | 112 (73-147) | **0.010** |
| Hemoglobin | 15.2 (14.3-15.9) | 15.4 (14.3-16.3) | 15.2 (13.9-15.7) | 0.234 |
| MCV | 96.1 (91.5-100.3) | 95.4 (93.2-97.8) | 97 (89.3-100.9) | 0.549 |
| INR | 1.10 (1.01-1.17) | 1.00 (1.00-1.10) | 1.10 (1.05-1.20) | **0.006** |
| Creatinine | 0.90 (0.81-1.04) | 0.88 (0.80-0.99) | 0.90 (0.81-1.04) | 0.562 |
| Glucose | 96 (88-101) | 99 (94-103) | 95 (88-100) | 0.278 |
| Albumin | 4.6 (4.3-4.8) | 4.6 (4.5-4.7) | 4.6 (4.2-4.8) | 0.772 |
| Bilirubin | 0.62 (0.41-0.90) | 0.50 (0.42-0.92) | 0.64 (0.41-0.88) | 0.677 |
| AST | 28.5 (24.3-32.0) | 22.5 (21.0-30.3) | 30.0 (26.3-33.5) | **0.005** |
| ALT | 24.0 (19.5-31.0) | 21.0 (17.0-27.5) | 25.5 (21.0-31.0) | 0.112 |
| Triglycerides | 118 (76-162) | 105 (83-172) | 118 (77-160) | 0.864 |
| Total cholesterol | 178 (155-194) | 179 (164-216) | 178 (146-190) | 0.412 |
| HDL | 43 (35-55) | 39 (33-54) | 45 (39-55) | 0.386 |
| LDL | 101 (83-122) | 109 (92-122) | 89 (82-122) | 0.413 |

**Supplementary Data 4**. Biochemical clinical data of HIV patients five years after completion of HCV therapy categorized by cirrhosis (LSM≥12.5kPa). **Statistics**: The values are expressed as the absolute number (percentage) and median (interquartile range). **Abbreviations**: MCV, mean corpuscular volume; INR, international normalized ratio; AST, aspartate transaminase; ALT, alanine aminotransferase; HDL, high-density hdllipoprotein; LDL, low-density lipoprotein.

| | **All patients** | **LSM<12.5kPa** | **LSM≥12.5kPa** | **p-value** |
|---|---|---|---|---|
| **No.** | 30 | 20 | 10 | |
| Neutrophils | 3675 (3033-4965) | 3675 (3025-4700) | 3945 (3133-4940) | 0.930 |
| Platelets ($10^3$) | 169 (140-214) | 171 (150-205) | 140 (105-227) | 0.403 |
| Hemoglobin | 15.8 (14.4-16.5) | 16.4 (15.3-17.0) | 14.7 (13.7-16.0) | **0.039** |
| MCV | 95.6 (91.9-100.2) | 94.6 (92.3-97.9) | 97.7 (92.1-104.8) | 0.322 |
| INR | 1.05 (1.00-1.10) | 1.02 (1.00-1.10) | 1.08 (1.00-1.10) | 0.507 |
| Creatinine | 0.90 (0.76-1.05) | 0.94 (0.86-1.04) | 0.80 (0.66-1.25) | 0.468 |
| Glucose | 92 (87-103) | 94 (87-102) | 90 (84-101) | 0.468 |
| Albumin | 4.6 (4.2-4.8) | 4.6 (4.2-4.8) | 4.6 (4.1-4.9) | 0.974 |
| Bilirubin | 0.56 (0.50-0.70) | 0.54 (0.49-0.69) | 0.60 (0.55-1.12) | 0.112 |
| AST | 26.0 (28.5-34.5) | 26.0 (18.0-31.0) | 24.5 (21.8-38.8) | 0.425 |
| ALT | 20.0 (14.0-36.0) | 25.0 (13.5-39.0) | 18.0 (15.5-23.8) | 0.713 |
| Triglycerides | 128 (83-181) | 128 (96-185) | 88 (58-162) | 0.232 |
| Total cholesterol | 176 (155-197) | 176 (162-195) | 181 (130-201) | 0.936 |
| HDL | 44 (40-57) | 43 (39-49) | 57 (42-62) | 0.543 |
| LDL | 93 (79-121) | 96 (81-118) | 86 (60-115) | 0.644 |

**Supplementary Data 5.** Proportion of main classes of identified lipids and lipid subclasses in each main class in people with HIV (PWH). **Abbreviations:** MG, monoglyceride; DG, diglyceride; TG, triglyceride; LPC, lysophosphatidylcholine; PC, phosphatidylcholine; PE, phosphatidylethanolamine; PI, phosphatidylinositol; PS, phosphatidylserine; SM, sphingomyelin; FA, fatty acid; FAHFA, fatty acyl ester of hydroxy fatty acid; LPE, lysophosphatidylethanolamine; PA, phosphatidic acid.

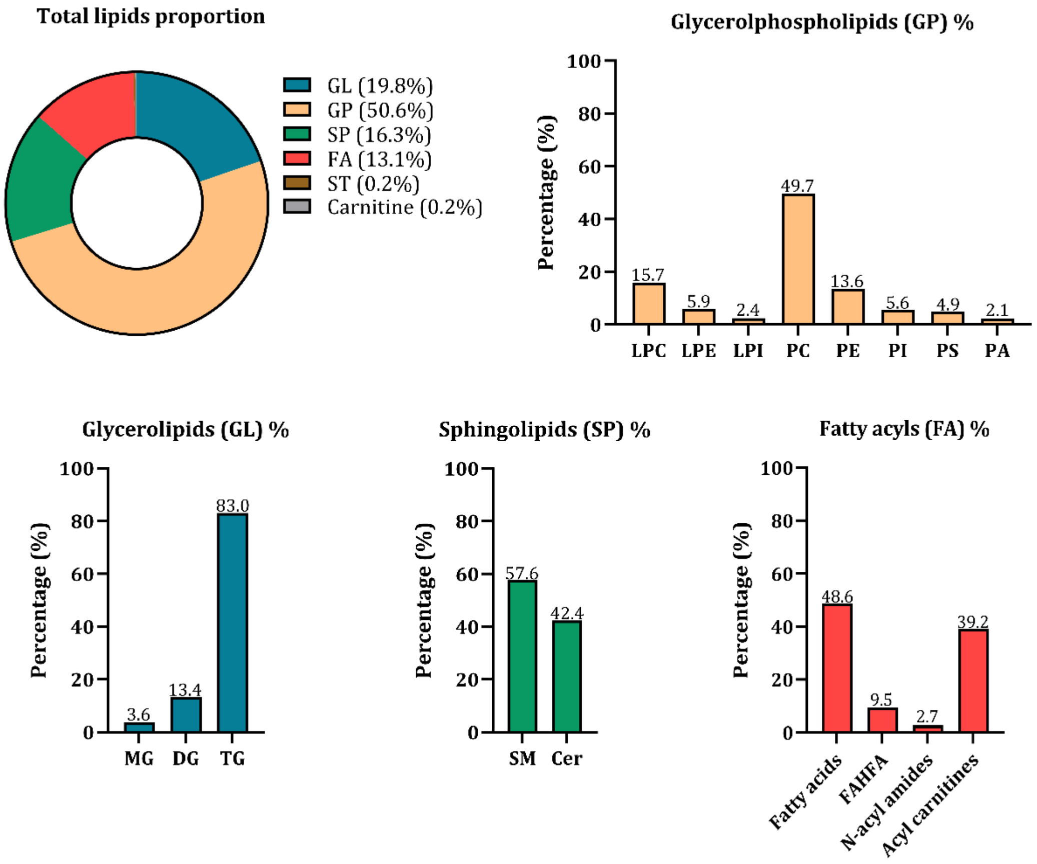

**Supplementary Data 6**. Association of individual metabolites with cirrhosis (LSM≥12.5kPa) at one year after completion of HCV therapy in PWH. **Statistics:** Association analysis was calculated by generalized linear models (GLM) with a gamma distribution (log-link) (dependent variable: plasma metabolites; independent variable: cirrhosis), adjusted by epidemiological characteristics (age, gender, body mass index, HCV treatment) using a forward stepwise method. *P*-values were adjusted by FDR correction for multiple comparisons (Benjamini and Hochberg).
**Abbreviations:** RT. retention time; aAMR. adjusted ratio of the arithmetic means; CI. confidence interval; p-value. level of significance; q-value. adjusted p-value by FDR correction; GC-MS. gas chromatography–mass spectrometry; LC-MS ESI+. liquid chromatography–mass spectrometry. positive electrospray ionization; LC-MS ESI+. liquid chromatography–mass spectrometry. negative electrospray ionization; LysoPC. lysophosphocoline; LysoPE. lysophosphatidylethanolamine;

| Feature | Technology | Mass | RT (min) | aAMR (CI95) | p | q |
|---|---|---|---|---|---|---|
| Methionine | CE-MS | 149.05 | 14.29 | 1.42 (1.22 - 1.64) | **<0.001** | **0.001** |
| Tyrosine | CE-MS | 181.08 | 14.99 | 1.27 (1.07 - 1.5) | **0.009** | **0.054** |
| Arginine | CE-MS | 174.11 | 10.49 | 1.66 (1.07 - 2.57) | **0.029** | **0.150** |
| Citrulline | CE-MS | 175.09 | 14.7 | 1.24 (1.07 - 1.44) | **0.007** | **0.053** |
| Cysteineglutathione_disulfide | CE-MS | 426.09 | 15.36 | 0.67 (0.47 - 0.96) | **0.034** | **0.150** |
| N-(1-Deoxy-1-fructosyl) methionine | CE-MS | 311.11 | 18.24 | 1.9 (1.24 - 2.9) | **0.005** | **0.053** |
| N-(1-Deoxy-1-fructosyl) phenylalanine | CE-MS | 327.13 | 18.29 | 1.7 (1.19 - 2.44) | **0.005** | **0.053** |
| Cer 18:0-O2/18:0 | LC-MS ESI+ | 567.56 | 11.71 | 1.03 (1.01 - 1.06) | **0.009** | **0.087** |
| Cer 18:0-O2/20:0 | LC-MS ESI+ | 595.59 | 11.98 | 0.98 (0.96 - 1) | **0.024** | **0.111** |
| CerPE 38:2-O2 | LC-MS ESI+ | 714.57 | 7.66 | 1.13 (1.01 - 1.27) | **0.044** | **0.121** |
| LPC 12:0/0:0 | LC-MS ESI+ | 439.27 | 1.22 | 1.15 (1.01 - 1.3) | **0.034** | **0.115** |
| LPC 17:0/0:0 | LC-MS ESI+ | 509.35 | 2.94 | 0.83 (0.7 - 0.99) | **0.041** | **0.119** |
| LPC 19:1/0:0 | LC-MS ESI+ | 535.36 | 3.08 | 0.9 (0.83 - 0.96) | **0.005** | **0.073** |
| LPC 20:3/0:0 | LC-MS ESI+ | 545.35 | 2.41 | 0.82 (0.68 - 0.98) | **0.038** | **0.116** |
| LPC 22:5/0:0 | LC-MS ESI+ | 569.35 | 2.24 | 0.82 (0.7 - 0.95) | **0.012** | **0.087** |
| LPE 0:0/16:0 | LC-MS ESI+ | 453.29 | 2.43 | 1.09 (1.02 - 1.16) | **0.019** | **0.111** |
| PC 12:0/16:0 | LC-MS ESI+ | 677.50 | 5.78 | 1.15 (1.05 - 1.27) | **0.006** | **0.078** |
| PC 14:0/16:1 | LC-MS ESI+ | 703.51 | 5.95 | 1.33 (1.16 - 1.54) | **<0.001** | **0.043** |
| PC 14:0/16:0 | LC-MS ESI+ | 705.53 | 6.99 | 1.59 (1.17 - 2.14) | **0.004** | **0.068** |
| PC 15:1/16:1 | LC-MS ESI+ | 715.51 | 7.96 | 1.1 (1.01 - 1.21) | **0.043** | **0.121** |
| PC 15:0/16:0 | LC-MS ESI+ | 719.55 | 7.64 | 1.32 (1.03 - 1.68) | **0.032** | **0.115** |
| PC 14:0/18:2 | LC-MS ESI+ | 729.53 | 6.24 | 1.3 (1.1 - 1.54) | **0.004** | **0.068** |
| PC 14:0/18:1 | LC-MS ESI+ | 731.55 | 7.23 | 1.71 (1.21 - 2.43) | **0.004** | **0.068** |
| PC 15:0/18:1 | LC-MS ESI+ | 745.56 | 8.03 | 1.2 (1.01 - 1.41) | **0.040** | **0.117** |
| PC 15:0/18:2 | LC-MS ESI+ | 743.55 | 6.89 | 1.26 (1.01 - 1.58) | **0.044** | **0.121** |
| PC 16:1/18:2 | LC-MS ESI+ | 755.55 | 6.43 | 1.28 (1.06 - 1.54) | **0.013** | **0.091** |
| PC 16:3/18:0 | LC-MS ESI+ | 755.55 | 6.86 | 1.3 (1.03 - 1.64) | **0.034** | **0.115** |
| PC 16:0/18:2 | LC-MS ESI+ | 757.56 | 7.64 | 1.32 (1.04 - 1.66) | **0.026** | **0.112** |

| | | | | | | |
|---|---|---|---|---|---|---|
| PC 18:2/18:2 | LC-MS ESI+ | 781.56 | 6.75 | 1.45 (1.16 - 1.82) | **0.002** | **0.068** |
| PC 18:1/18:2 | LC-MS ESI+ | 783.58 | 7.86 | 1.34 (1.12 - 1.6) | **0.003** | **0.068** |
| PC 18:0/18:2 | LC-MS ESI+ | 785.59 | 9.65 | 1.43 (1.04 - 1.96) | **0.031** | **0.115** |
| PC 18:1/20:2 | LC-MS ESI+ | 811.61 | 9.8 | 1.24 (1.08 - 1.42) | **0.003** | **0.068** |
| PC 20:5/21:0 | LC-MS ESI+ | 849.63 | 7.63 | 1.29 (1.04 - 1.6) | **0.023** | **0.111** |
| PC 20:4/22:4 | LC-MS ESI+ | 857.59 | 7.64 | 1.28 (1.07 - 1.53) | **0.010** | **0.087** |
| PC 33:3 | LC-MS ESI+ | 741.54 | 7.64 | 1.33 (1.04 - 1.7) | **0.028** | **0.112** |
| PC 39:2 | LC-MS ESI+ | 827.64 | 9.65 | 1.42 (1.05 - 1.91) | **0.028** | **0.112** |
| PC O-14:0/18:2 | LC-MS ESI+ | 715.55 | 7.23 | 1.26 (1.09 - 1.47) | **0.004** | **0.068** |
| PC O-16:0/14:1 | LC-MS ESI+ | 689.53 | 7.77 | 1.2 (1.02 - 1.41) | **0.036** | **0.115** |
| PC O-16:0/20:4 | LC-MS ESI+ | 767.58 | 8.56 | 1.26 (1.04 - 1.54) | **0.022** | **0.111** |
| PC O-16:1/18:2 | LC-MS ESI+ | 741.57 | 8.52 | 1.32 (1.07 - 1.62) | **0.011** | **0.087** |
| PC O-18:1/18:2 | LC-MS ESI+ | 785.59 | 9.65 | 1.46 (1.07 - 2) | **0.022** | **0.111** |
| PE 16:0/18:2 | LC-MS ESI+ | 715.52 | 7.96 | 1.1 (1 - 1.21) | **0.047** | **0.127** |
| PE 16:0/20:5 | LC-MS ESI+ | 737.50 | 7.22 | 1.19 (1.04 - 1.36) | **0.014** | **0.092** |
| PE 18:0/18:2 | LC-MS ESI+ | 743.55 | 10.12 | 1.18 (1.01 - 1.36) | **0.036** | **0.115** |
| PE 18:0/20:4 | LC-MS ESI+ | 767.55 | 9.66 | 1.46 (1.06 - 1.99) | **0.023** | **0.111** |
| PE 18:1/18:2 | LC-MS ESI+ | 741.54 | 8.16 | 1.17 (1.02 - 1.34) | **0.029** | **0.112** |
| PE 18:2/20:1 | LC-MS ESI+ | 769.56 | 7.85 | 1.25 (1.09 - 1.43) | **0.002** | **0.068** |
| PE 18:2/20:0 | LC-MS ESI+ | 771.58 | 9.65 | 1.28 (1.05 - 1.57) | **0.018** | **0.111** |
| PE 36:4 | LC-MS ESI+ | 739.52 | 7.64 | 1.29 (1.04 - 1.61) | **0.026** | **0.112** |
| PE O-16:1/20:4 | LC-MS ESI+ | 723.52 | 8.52 | 1.31 (1.07 - 1.6) | **0.010** | **0.087** |
| PE O-18:0/20:4 | LC-MS ESI+ | 753.57 | 11.3 | 1.15 (1.02 - 1.29) | **0.026** | **0.112** |
| PE O-18:2/18:2 | LC-MS ESI+ | 725.54 | 9.23 | 1.08 (1.02 - 1.15) | **0.009** | **0.087** |
| PE O-18:2/20:4 | LC-MS ESI+ | 749.54 | 8.56 | 1.28 (1.05 - 1.56) | **0.019** | **0.111** |
| PE O-38:5 | LC-MS ESI+ | 751.55 | 7.64 | 1.28 (1.08 - 1.52) | **0.007** | **0.078** |
| PE O-38:4 | LC-MS ESI+ | 753.57 | 11.3 | 1.15 (1.02 - 1.28) | **0.022** | **0.111** |
| PI 18:0/18:2 | LC-MS ESI+ | 862.55 | 7.64 | 1.19 (1.02 - 1.38) | **0.031** | **0.115** |
| PS 41:6 | LC-MS ESI+ | 849.56 | 9.65 | 1.22 (1.05 - 1.41) | **0.012** | **0.087** |
| SM 18:0; O2/16:0 | LC-MS ESI+ | 704.58 | 7.59 | 1.27 (1.02 - 1.58) | **0.036** | **0.115** |
| SM 18:1; O2/18:0 | LC-MS ESI+ | 730.60 | 7.64 | 1.23 (1.02 - 1.48) | **0.037** | **0.115** |
| SM 18:1; O2/25:0 | LC-MS ESI+ | 828.71 | 12.65 | 0.99 (0.98 - 1) | **0.037** | **0.115** |
| TG 52:5 | LC-MS ESI+ | 852.72 | 13.08 | 0.99 (0.97 - 1) | **0.039** | **0.117** |
| TG 53:5 | LC-MS ESI+ | 866.73 | 13.24 | 0.98 (0.96 - 0.99) | **0.004** | **0.068** |
| TG 54:0 | LC-MS ESI+ | 890.83 | 16.64 | 0.98 (0.97 - 1) | **0.020** | **0.111** |
| TG 56:7 | LC-MS ESI+ | 902.74 | 13.37 | 0.98 (0.96 - 0.99) | **0.011** | **0.087** |
| Cer 18:1-O2/16:1 | LC-MS ESI- | 535.50 | 7.62 | 1.1 (1.03 - 1.19) | **0.011** | **0.058** |
| Cer 20:0-O2/24:0 | LC-MS ESI- | 679.68 | 12.88 | 0.98 (0.96 - 0.99) | **0.009** | **0.058** |
| FA 20:0 | LC-MS ESI- | 312.30 | 4.96 | 0.95 (0.91 - 0.98) | **0.006** | **0.053** |
| FA 34:0 | LC-MS ESI- | 508.52 | 12.85 | 1.02 (1 - 1.03) | **0.047** | **0.149** |
| LPI 18:2/0:0 | LC-MS ESI- | 596.30 | 1.65 | 1.28 (1.02 - 1.6) | **0.035** | **0.119** |
| PC 16:0/16:1 | LC-MS ESI- | 731.55 | 7.21 | 1.34 (1.08 - 1.67) | **0.011** | **0.058** |
| PC 39:6 | LC-MS ESI- | 819.59 | 7.61 | 1.23 (1.04 - 1.45) | **0.021** | **0.099** |

| | | | | | | |
|---|---|---|---|---|---|---|
| PC O-18:0/12:0 | LC-MS ESI- | 691.55 | 8.21 | 1.12 (1.01 - 1.24) | **0.033** | **0.119** |
| PE O-18:1/18:2 | LC-MS ESI- | 727.55 | 11.32 | 1.21 (1.08 - 1.36) | **0.002** | **0.053** |
| PI 16:0/18:2 | LC-MS ESI- | 834.53 | 6 | 1.5 (1.15 - 1.95) | **0.004** | **0.053** |
| PS 18:2/20:0 | LC-MS ESI- | 815.57 | 6.42 | 1.25 (1.07 - 1.46) | **0.006** | **0.053** |
| PS 18:1/20:0 | LC-MS ESI- | 817.58 | 7.61 | 1.25 (1.04 - 1.52) | **0.024** | **0.102** |
| PS 18:2/22:1 | LC-MS ESI- | 841.58 | 6.73 | 1.41 (1.15 - 1.73) | **0.002** | **0.053** |
| PS 18:2/22:0 | LC-MS ESI- | 843.60 | 7.83 | 1.26 (1.08 - 1.48) | **0.005** | **0.053** |
| SM 16:1;O2/24:0 | LC-MS ESI- | 808.64 | 9.62 | 1.33 (1.04 - 1.71) | **0.028** | **0.111** |
| SM 18:3;O2/20:0 | LC-MS ESI- | 754.60 | 7.21 | 1.35 (1.08 - 1.69) | **0.010** | **0.058** |

**Supplementary Data 7.** Chemical composition of glycerophospholipids species associated with cirrhosis (LSM≥12.5kPa) in people with HIV (PWH). (**A**) One year after completion of HCV therapy. (**B**) Five years after completion of HCV therapy. **Abbreviations:** PC, phosphatidylcholine; PE, phosphatidylethanolamine; PI, phosphatidylinositol; PS, phosphatidylserine.

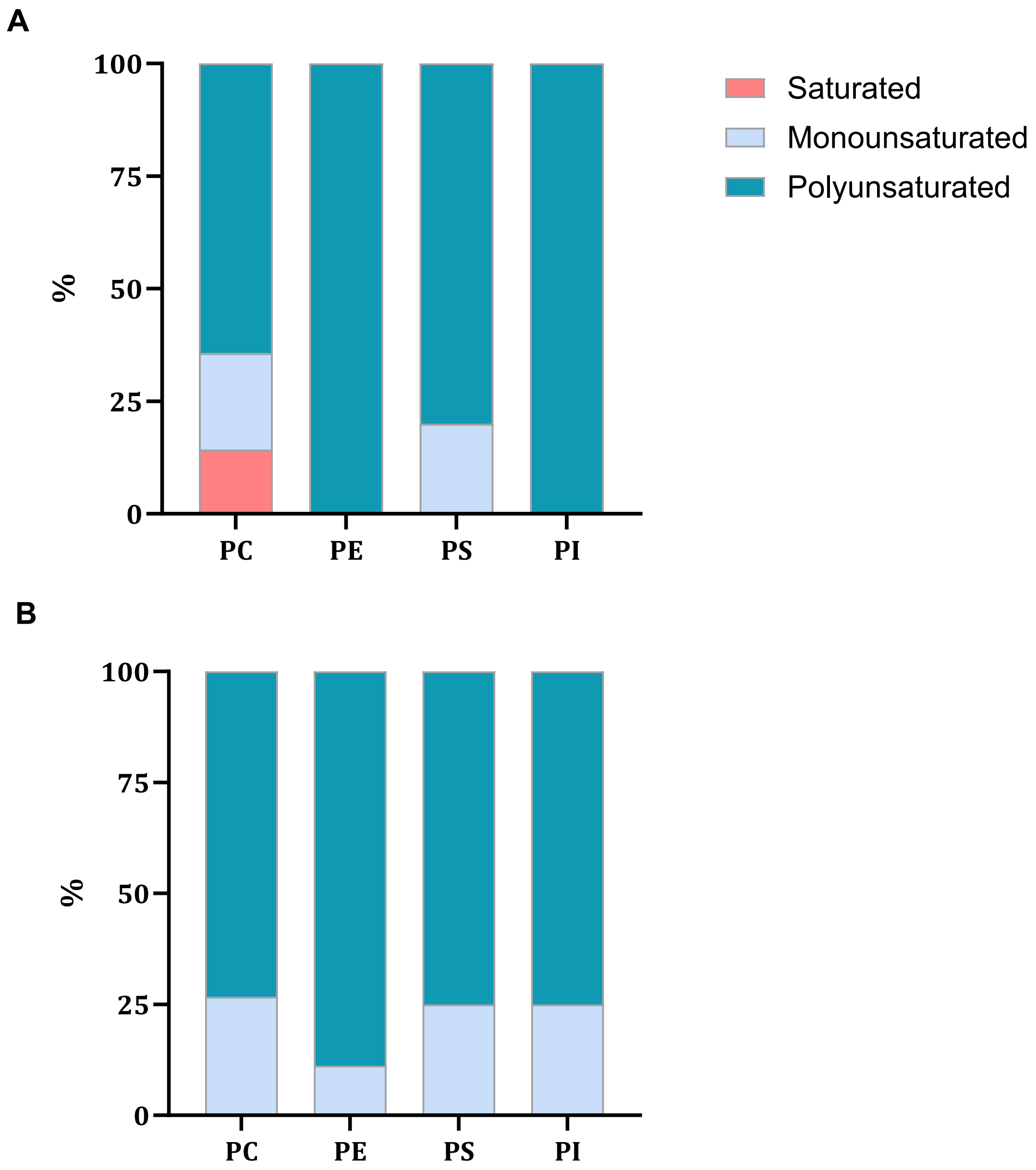

**Supplementary Data 8**. Association of individual metabolites with cirrhosis (LSM≥12.5kPa) five years after completion of HCV therapy in PWH. **Statistics:** Association analysis was calculated by generalized linear models (GLM) with a gamma distribution (log-link) (dependent variable: plasma metabolites; independent variable: cirrhosis), adjusted by epidemiological characteristics (age, gender, body mass index, HCV treatment). *P*-values were adjusted by FDR correction for multiple comparisons (Benjamini and Hochberg).

**Abbreviations:** RT. retention time; aAMR. adjusted ratio of the arithmetic means; CI. confidence interval; p-value. level of significance; q-value. adjusted p-value by FDR correction; GC-MS. gas chromatography–mass spectrometry; LC-MS ESI+. liquid chromatography–mass spectrometry. positive electrospray ionization; LC-MS ESI+. liquid chromatography–mass spectrometry. negative electrospray ionization; LysoPC. lysophosphocoline; LysoPE. lysophosphatidylethanolamine;

| **Feature** | **Technology** | **Mass** | **RT (min)** | **aAMR (CI95)** | **p** | **q** |
|---|---|---|---|---|---|---|
| L-Valine | CE-MS | 117.08 | 13.59 | 0.79 (0.68 - 0.90) | **0.002** | **0.022** |
| Hydroxyproline | CE-MS | 131.06 | 15.73 | 1.76 (1.24 - 2.51) | **0.004** | **0.030** |
| L-Lysine | CE-MS | 146.11 | 10.18 | 0.79 (0.67 - 0.94) | **0.010** | **0.039** |
| L-Tryptophan | CE-MS | 204.09 | 14.61 | 0.82 (0.68 - 0.98) | **0.037** | **0.097** |
| Norvaline | CE-MS | 117.08 | 13.59 | 0.79 (0.68 - 0.90) | **0.002** | **0.022** |
| L-Alloisoleucine | CE-MS | 131.09 | 13.89 | 0.83 (0.7 - 0.99) | **0.044** | **0.097** |
| L-Aspartic_acid | CE-MS | 133.04 | 15.14 | 0.76 (0.59 - 0.99) | **0.049** | **0.099** |
| Arginine | CE-MS | 174.11 | 10.49 | 2.51 (1.32 - 4.75) | **0.009** | **0.039** |
| Citrulline | CE-MS | 175.10 | 14.7 | 1.29 (1.07 - 1.54) | **0.012** | **0.040** |
| D-Alanyl-D-Valine | CE-MS | 188.12 | 12.53 | 0.72 (0.57 - 0.91) | **0.009** | **0.039** |
| Homo-L-arginine | CE-MS | 188.13 | 10.89 | 0.7 (0.56 - 0.89) | **0.007** | **0.039** |
| Propionyl-L-carnitine | CE-MS | 217.13 | 12.86 | 1.33 (1.02 - 1.75) | **0.045** | **0.097** |
| (R)-Pantetheine | CE-MS | 278.13 | 13.6 | 0.64 (0.5 - 0.82) | **0.001** | **0.022** |
| Cysteineglutathione_disulfide | CE-MS | 426.09 | 15.36 | 0.49 (0.26 - 0.91) | **0.032** | **0.095** |
| Pyroglutamine | CE-MS | 128.06 | 12.29 | 1.35 (1.03 - 1.76) | **0.039** | **0.097** |
| CAR 10:1 | LC-MS ESI+ | 313.23 | 0.88 | 1.42 (1.03 - 1.95) | **0.040** | **0.191** |
| CAR 9:0 | LC-MS ESI+ | 301.23 | 0.85 | 1.49 (1.15 - 1.93) | **0.005** | **0.102** |
| Cer 18:0-O2/24:0 | LC-MS ESI+ | 651.65 | 12.56 | 0.97 (0.95 – 1.00) | **0.026** | **0.173** |
| Cer 18:1-O2/25:0 | LC-MS ESI+ | 663.65 | 12.55 | 0.97 (0.95 – 1.00) | **0.040** | **0.191** |
| LPC 20:4/0:0 | LC-MS ESI+ | 543.33 | 2.07 | 0.76 (0.62 - 0.94) | **0.015** | **0.138** |
| LPC 22:1/0:0 | LC-MS ESI+ | 577.41 | 4.2 | 1.13 (1.04 - 1.22) | **0.006** | **0.102** |
| LPE 18:2/0:0 | LC-MS ESI+ | 477.28 | 2.22 | 1.25 (1.02 - 1.54) | **0.045** | **0.191** |
| LPE 20:4/0:0 | LC-MS ESI+ | 501.28 | 2.12 | 1.28 (1.03 - 1.6) | **0.037** | **0.191** |
| LPE 22:6/0:0 | LC-MS ESI+ | 525.29 | 2.07 | 0.81 (0.66 - 0.98) | **0.038** | **0.191** |
| PC 15:1/16:1 | LC-MS ESI+ | 715.51 | 7.96 | 1.2 (1.07 - 1.35) | **0.004** | **0.102** |
| PC 16:1/18:2 | LC-MS ESI+ | 755.55 | 6.43 | 1.43 (1.11 - 1.85) | **0.010** | **0.138** |
| PC 18:1/18:2 | LC-MS ESI+ | 783.58 | 7.86 | 1.56 (1.23 - 1.99) | **0.001** | **0.091** |

| | | | | | | |
|---|---|---|---|---|---|---|
| PC 18:1/18:1 | LC-MS ESI+ | 785.59 | 9.3 | 1.44 (1.1 - 1.88) | **0.013** | **0.138** |
| PC 18:1/20:2 | LC-MS ESI+ | 811.61 | 9.8 | 1.32 (1.07 - 1.62) | **0.013** | **0.138** |
| PC 18:1/20:1 | LC-MS ESI+ | 813.62 | 11.5 | 1.19 (1.03 - 1.37) | **0.023** | **0.173** |
| PC 18:2/18:2 | LC-MS ESI+ | 781.56 | 6.75 | 1.81 (1.37 - 2.4) | **<0.001** | **0.042** |
| PC 38:1 | LC-MS ESI+ | 815.64 | 11.87 | 1.04 (1.01 - 1.07) | **0.016** | **0.138** |
| PC 40:6 | LC-MS ESI+ | 833.59 | 7.84 | 1.19 (1.01 - 1.41) | **0.046** | **0.191** |
| PC O-16:1/18:2 | LC-MS ESI+ | 741.57 | 8.52 | 1.37 (1.02 - 1.85) | **0.047** | **0.191** |
| PC O-18:1/22:6 | LC-MS ESI+ | 817.59 | 8.34 | 1.24 (1.05 - 1.45) | **0.016** | **0.138** |
| PC O-40:8 | LC-MS ESI+ | 815.58 | 7.32 | 1.27 (1.05 - 1.55) | **0.022** | **0.173** |
| PC O-42:5 | LC-MS ESI+ | 863.64 | 11.54 | 1.12 (1.02 - 1.23) | **0.032** | **0.188** |
| PE 16:0/18:1 | LC-MS ESI+ | 717.53 | 9.38 | 1.13 (1.03 - 1.23) | **0.016** | **0.138** |
| PE 16:0/18:2 | LC-MS ESI+ | 715.52 | 7.96 | 1.2 (1.07 - 1.35) | **0.005** | **0.102** |
| PE 18:0/18:2 | LC-MS ESI+ | 743.55 | 10.12 | 1.27 (1.09 - 1.49) | **0.006** | **0.102** |
| PE 18:1/18:2 | LC-MS ESI+ | 741.53 | 8.16 | 1.3 (1.06 - 1.59) | **0.016** | **0.138** |
| PE 18:2/20:1 | LC-MS ESI+ | 769.56 | 7.85 | 1.42 (1.16 - 1.74) | **0.002** | **0.102** |
| PE O-16:1/20:5 | LC-MS ESI+ | 721.50 | 7.64 | 1.24 (1.02 - 1.51) | **0.044** | **0.191** |
| PE O-18:0/20:4 | LC-MS ESI+ | 753.57 | 11.3 | 1.17 (1.03 - 1.34) | **0.028** | **0.173** |
| PE O-38:4 | LC-MS ESI+ | 753.57 | 11.3 | 1.17 (1.03 - 1.33) | **0.027** | **0.173** |
| PI 16:0/22:5 | LC-MS ESI+ | 884.54 | 6.05 | 1.11 (1.02 - 1.22) | **0.026** | **0.173** |
| TG 14:0/13:1/18:1 | LC-MS ESI+ | 760.66 | 13.01 | 0.98 (0.97 – 1.00) | **0.046** | **0.191** |
| TG 42:0* | LC-MS ESI+ | 722.64 | 13.09 | 0.98 (0.96 – 1.00) | **0.039** | **0.191** |
| TG 49:1** | LC-MS ESI+ | 818.76 | 14.15 | 0.97 (0.95 – 1.00) | **0.047** | **0.191** |
| TG 49:2*** | LC-MS ESI+ | 816.72 | 13.74 | 0.98 (0.96 – 1.00) | **0.028** | **0.173** |
| TG 56:6-OH | LC-MS ESI+ | 922.76 | 13.94 | 1.03 (1.01 - 1.05) | **0.006** | **0.102** |
| TG 56:7 | LC-MS ESI+ | 902.73 | 13.37 | 0.98 (0.96 - 0.99) | **0.011** | **0.138** |
| Cer 20:0-O2/24:0 | LC-MS ESI- | 679.68 | 12.88 | 0.97 (0.94 – 1.00) | **0.037** | **0.114** |
| DG 38:4 | LC-MS ESI- | 644.54 | 11.95 | 1.03 (1.01 - 1.06) | **0.024** | **0.093** |
| FA_24:0 | LC-MS ESI- | 368.37 | 7.17 | 1.09 (1 - 1.17) | **0.050** | **0.134** |
| FAHFA 16:0/18:2 | LC-MS ESI- | 534.46 | 3.37 | 1.42 (1.08 - 1.86) | **0.018** | **0.092** |
| LPI 18:0/0:0 | LC-MS ESI- | 600.33 | 2.79 | 1.27 (1.02 - 1.57) | **0.041** | **0.118** |
| LPI 18:2/0:0 | LC-MS ESI- | 596.30 | 1.65 | 1.41 (1.17 - 1.69) | **0.001** | **0.016** |
| PC 36:5 | LC-MS ESI- | 779.55 | 5.98 | 1.38 (1.09 - 1.76) | **0.014** | **0.092** |
| PC O-18:0/18:2 | LC-MS ESI- | 771.62 | 11.52 | 1.23 (1.05 - 1.44) | **0.019** | **0.092** |
| PE O-18:1/18:2 | LC-MS ESI- | 727.55 | 11.32 | 1.24 (1.07 - 1.45) | **0.010** | **0.090** |
| PI 16:0/18:1 | LC-MS ESI- | 836.54 | 6.83 | 1.38 (1.07 - 1.79) | **0.020** | **0.092** |
| PI 16:0/18:2 | LC-MS ESI- | 834.52 | 6 | 1.39 (1.06 - 1.82) | **0.024** | **0.093** |
| PI 38:6 | LC-MS ESI- | 882.52 | 5.73 | 1.36 (1.03 - 1.79) | **0.037** | **0.114** |
| PS 18:2/20:0 | LC-MS ESI- | 815.57 | 6.42 | 1.34 (1.08 - 1.66) | **0.013** | **0.092** |
| PS 18:2/22:0 | LC-MS ESI- | 843.60 | 7.83 | 1.57 (1.27 - 1.96) | **0.000** | **0.008** |
| PS 18:2/22:1 | LC-MS ESI- | 841.58 | 6.73 | 1.75 (1.35 - 2.28) | **0.000** | **0.008** |
| PS 22:0/18:1 | LC-MS ESI- | 845.61 | 9.27 | 1.49 (1.14 - 1.94) | **0.007** | **0.077** |
| SM 42:4-O2 | LC-MS ESI- | 808.64 | 9.62 | 1.39 (1.04 - 1.87) | **0.036** | **0.114** |

**Notes**: Since a higher chromatographic resolution was not possible, different chain isomers with the same overall composition were detected.

* TG 10:0/14:0/18:0 // TG 13:0/14:0/15:0 // TG 12:0/14:0/16:0 //TG 14:0/14:0/14:0 //TG 12:0/12:0/18:0 //TG 10:0/16:0/16:0 //TG 10:0/14:0/18:0

** TG 15:0/16:0/18:1 // TG 15:0/18:0/16:1 // TG 16:0/16:0/17:1 // TG 16:0/17:0/16:1

*** TG 14:0/17:1/18:1 // TG 15:0/16:0/18:2 // TG 15:0/16:1/18:1 // TG 15:0/17:1/17:1 // TG 16:0/16:1/17:1 // TG 16:1/16:1/17:0
